# Highly multiplexed spatial analysis identifies tissue-resident memory T cells as drivers of ulcerative and immune checkpoint inhibitor induced colitis

**DOI:** 10.1101/2023.04.25.23289056

**Authors:** Mick J.M. van Eijs, José J.M. ter Linde, Matthijs J.D. Baars, Mojtaba Amini, Miangela M. Laclé, Eelco C. Brand, Eveline M. Delemarre, Julia Drylewicz, Stefan Nierkens, Rik J. Verheijden, Bas Oldenburg, Yvonne Vercoulen, Karijn P.M. Suijkerbuijk, Femke van Wijk

## Abstract

Colitis is a prevalent adverse event associated with immune checkpoint inhibitor (ICI) therapy with similarities to inflammatory bowel disease. Incomplete mechanistic understanding of ICI-colitis curtails evidence-based treatment. Given the often-overlooked connection between tissue architecture and mucosal immune cell function, we here applied imaging mass cytometry (IMC) to gain spatial proteomic insight in ICI-colitis in comparison to ulcerative colitis (UC). Using a cell segmentation pipeline that simultaneously utilizes high-resolution nuclear imaging and high-multiplexity IMC, we show that CD8^+^ T cells are significantly more abundant (and dominant) in anti-PD-1 +/-anti-CTLA-4-induced colitis compared to anti-CTLA-4-induced colitis and UC. We identified activated, cycling CD8^+^ tissue-resident memory T (T_RM_) cells at the lamina propria-epithelial interface as drivers of cytotoxicity in ICI-colitis and UC. Moreover, we found that combined ICI-induced colitis featured highest granzyme B levels both in tissue and serum. Together, these data reinforce CD8^+^ T_RM_ cells as potentially targetable drivers of ICI-colitis.

## Introduction

Treatment of various advanced malignancies has considerably improved with the introduction of immune checkpoint inhibitors (ICIs). However, a major downside associated with ICI therapy remains its large variety of immune-related adverse events (irAEs).^1^ ICI-colitis is among the most prevalent irAEs at approximately 16% with combined anti-cytotoxic T-lymphocyte-associated protein 4 (αCTLA-4) and anti-programmed death 1 (αPD-1) therapy (cICI) and is potentially lethal if not adequately treated.^2,3^ Depending on severity, ICI-colitis requires ICI discontinuation, administration of high-dose systemic steroids or selective immunosuppression in steroid-refractory cases.^4,5^ Step-up to selective immunosuppression is largely expert-opinion driven and mostly based on experience in the treatment of patients with inflammatory bowel disease (IBD). Infliximab and vedolizumab are therefore frequently used.^6^ Notwithstanding clinical parallels between ICI-colitis and IBD, both conditions should be considered separate diseases. This is underscored by lesser chronicity in histopathology, superior biological response rate and shorter time-to-response in ICI-colitis than IBD.^6–9^

ICI-colitis can be subdivided based on the specific ICI regimen, i.e., αCTLA-4 monotherapy, αPD-1 monotherapy or combined (c)ICI. Clinically, time-to-onset of αPD-1 colitis is usually longer than for αCTLA-4-based regimes, while response to steroids is better for αPD-1 colitis than for colitis after αCTLA-4 +/-αPD-1.^10,11^ Immunohistochemistry (IHC) studies have characterized αCTLA-4 colitis as CD4^+^ T lymphocytic disease with increased interferon (IFN)-γ; and interleukin (IL)-17 responses, intra-epithelial neutrophils and erosions.^12,13^ In contrast, αPD-1 colitis has been associated with CD8^+^ intra-epithelial lymphocytosis,^13^ although another study that also reported overlapping histopathology between ulcerative, Crohn’s and αCTLA-4 colitis found lower CD8 staining scores for αPD-1 than for αCTLA-4 colitis.^9^ With the advent of single-cell RNA-sequencing (scRNA-seq), enabling highly comprehensive phenotyping, the role of CD8^+^ tissue-resident memory (T_RM_) cells could be explored in more detail.^14–17^ Some reports indicate a relative decrease of colonic CD8^+^ T_RM_ cells in ICI-colitis compared to ICI-treated patients without colitis,^14,16^ possibly due to either an influx of circulating effector T cells, or transformation of resting T_RM_ into inflammatory effectors with the loss of residency markers. CD8^+^ T_RM_ cells are also considered pro-inflammatory actors in IBD,^18^ although, similar to ICI-colitis, a relative decrease in CD8^+^ T_RM_ cells in active IBD versus healthy control tissue has been found.^19,20^

Interestingly, intra-epithelial CD8^+^ T_RM_ cells have been shown to transcriptionally acquire an innate pro-inflammatory profile in inflammatory sites of Crohn’s ileitis,^21^ illustrating the relevance of epithelial-lamina propria localization in relation to cell function. Yet, transcriptomic studies on tissue in ICI-colitis inherently lack architectural insight. Incomplete pathophysiologic understanding of causes for the clinically relevant differences observed between UC and ICI-colitis after different ICI regimens prompts a direct in-depth comparison among these groups. Using imaging mass cytometry (IMC), we aimed to integrally characterize immune cell, particularly T cell, infiltrates in different types of ICI-colitis and UC, both spatially and highly multiplexed at the same time. We adopted a novel IMC analysis approach to reliably segment and annotate single cells and we directly compared UC to different subtypes of ICI-colitis. Our study identifies CD8^+^ T cells as the dominant immune cell population in αPD-1 and cICI-colitis and shows that activated, cycling CD8^+^ T_RM_ cells residing close to the epithelial border are drivers of cytotoxicity across different types of ICI- and ulcerative colitis.

## Results

### Study design and population

Colon biopsies from 18 patients with ICI-colitis (4 αCTLA-4 monotherapy, 5 αPD-1 monotherapy, 9 cICI) and 5 patients with UC were included for sequential DAPI nuclear imaging and IMC (‘IMC cohort’; **Fig. 1A,B, Supplementary Table 1**). Nine of 23 patients (39%) were female, median age was 67 years (p25–p75, 51–72) and most patients with ICI-colitis were treated for melanoma (78%). The majority of patients (61%) was steroid-naïve upon endoscopy (median 0 days of steroid use; p25–p75, 0–1). Three of five patients with UC and all ICI-colitis patients underwent endoscopy for new-onset disease. In addition, we included serum samples from 80 ICI-treated patients, of whom 39 developed clinically relevant irAEs including 14 (36%) with colitis, for multiplexed proteomics. These serum samples were collected at baseline and upon onset of irAEs or ±6 weeks after treatment initiation for irAE-free patients (‘Serum cohort’; **Fig. 1A, Supplementary Table 1**). Four cICI-treated patients included in the IMC cohort were also part of this Serum cohort. Finally, we reanalyzed previously published colonic CD3^+^ scRNA-seq data (GSE144469) to validate main findings from the IMC cohort (**Fig. 1A**).^14^

**Figure 1.**
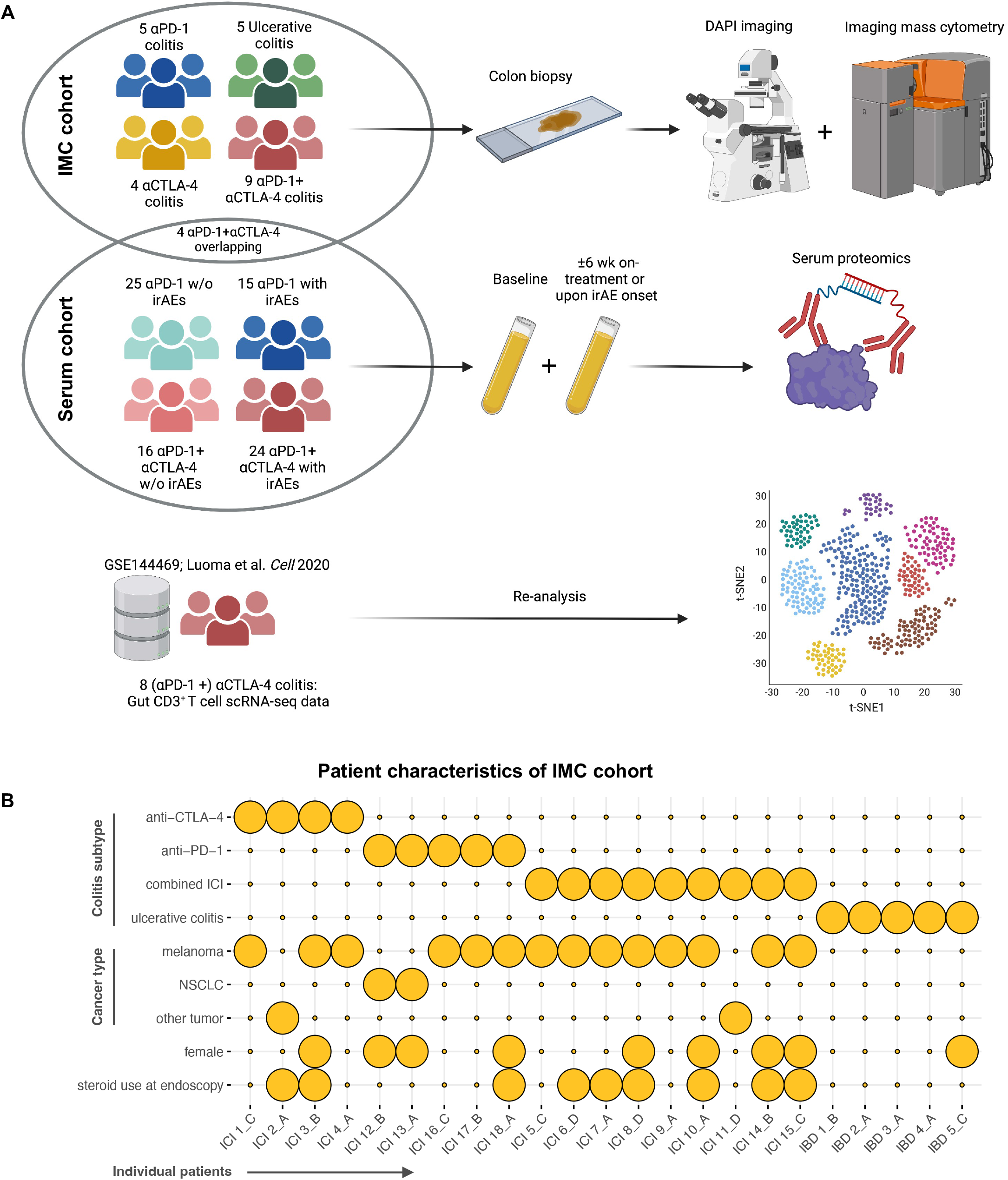
Study design and participant characteristics. **A**. All analyses were performed with data from (1) combined DAPI-imaging and imaging mass cytometry (IMC) of colitis tissue (n=23); (2) paired serum samples from baseline and either 6 weeks after αPD-1 monotherapy or combined αCTLA-4 + αPD-1 treatment initiation, or upon irAEs (n=80); and (3) previously published colonic T cell single-cell RNA-sequencing (scRNA-seq) data from patients with colitis after αCTLA-4 with or without αPD-1 treatment (n=8).^14^ Created with BioRender.com. **B**. Characteristics of patients in the IMC cohort. Characteristics that apply to a given patient are indicated by yellow spheres.

First, we explored to what extent histopathologic and endoscopic disease severity indices, commonly used in UC and increasingly in ICI-colitis, were correlated. Within the IMC cohort, a modest association was observed between both the Geboes score for ‘erosion/ulceration’ and the Robarts Histopathology Index (RHI) with the Mayo endoscopic score. The Geboes score for ‘chronic inflammatory infiltrate’ showed a moderate correlation with symptom duration (**Supplementary Fig. 1A-C**). All tissue samples histologically featured inflammation in line with the diagnoses of ICI-or ulcerative colitis and were therefore included for further analysis.

### CD8^+^ T cells are the dominant immune cell population in αPD-1 and combined-ICI-colitis

We acquired hematoxylin & eosin (H&E)-stained, DAPI and IMC images for all 23 patients with colitis in the IMC cohort (**Fig. 2A**). Using the MATISSE cell segmentation pipeline that simultaneously takes advantage of high-resolution DAPI nuclear imaging in combination with high-multiplexity IMC,^22,23^ we identified 215,293 single cells across all patient samples. After removal of cells from excluded tissue regions (including artefacts, lymphoid follicles and submucosa), 184,975 single cells remained for data normalization, scaling and lineage determination. These cells were used to generate pseudo-color cell-type annotated images as shown in **Fig. 2A**. Due to imaging-inherent overlap of membrane signal between neighboring cells, especially in densely populated infiltrates as seen in colitis, we deliberately developed a *supervised* cell type annotation approach. A comparable strategy previously achieved reliable annotation of segmented intestinal immune cells.^24^ Indeed, a uniform manifold approximation and projection (UMAP) visualization overlaid with supervised annotation labels confirmed that unsupervised clustering alone would probably insufficiently distinguish biologically distinct cell clusters (**Fig. 2B**).

**Figure 2.**
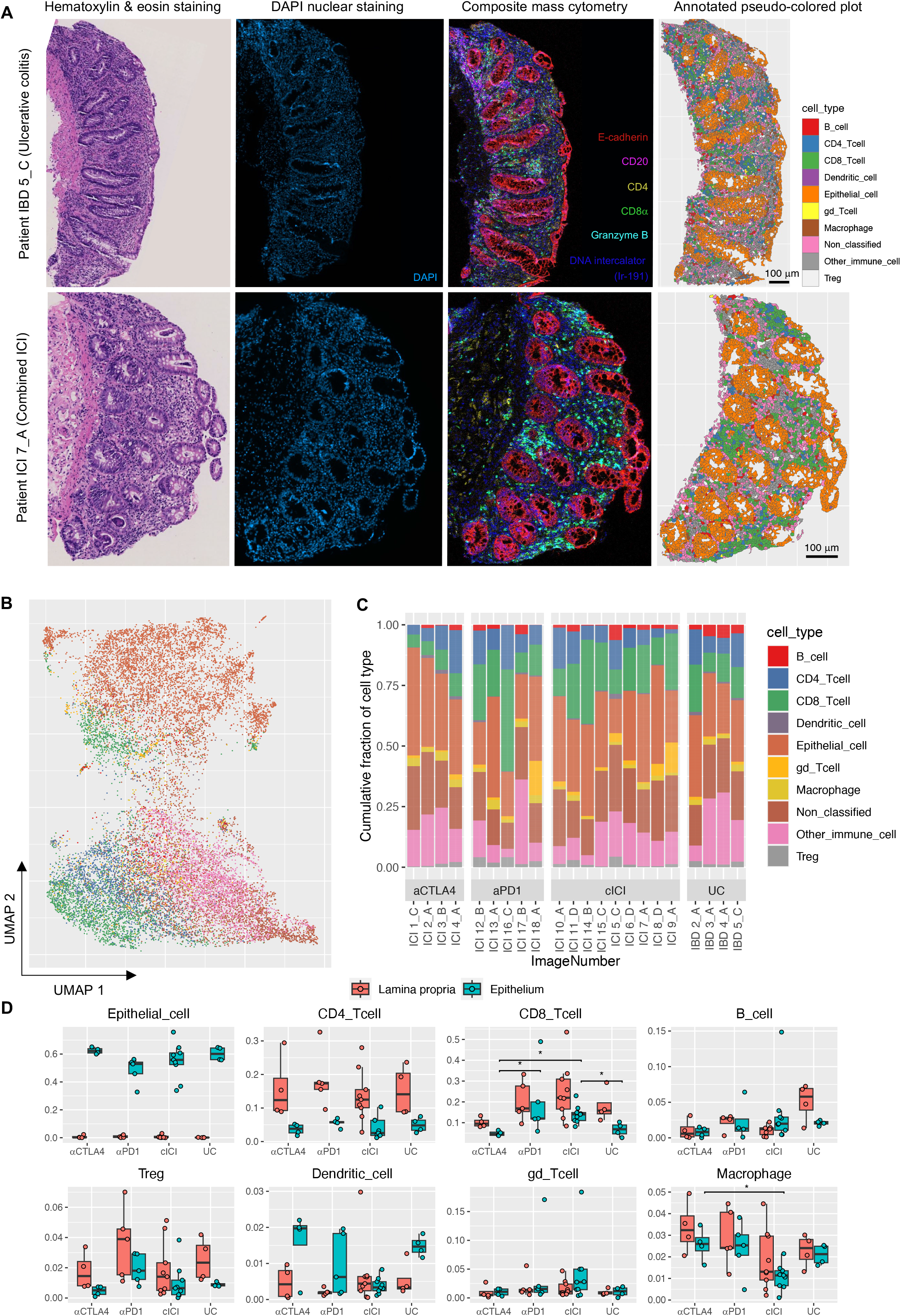
Cell type distribution across individual samples after cell segmentation and supervised lineage determination. **(A)**Representative sample images from patients with ulcerative colitis (top) and combined αCTLA-4 and αPD-1 colitis (bottom). Shown are hematoxylin & eosin stained images (adjacent serial section) used for histopathologic annotations, DAPI nuclear images, pseudo-color composite mass cytometry images and finally representations of extracted single-cell events after data normalization, scaling and lineage determination, pseudo-colored by cell type. **(B)**Uniform Manifold Approximation and Projection (UMAP) visualization of a random subset comprising 10% of cells across all patients with IMC data of sufficient quality (n=22), colored by assigned cell type label with color-coding as in (**C**). **(C)**Stacked bar graph representing fractions of cell types across all patient samples. cICI denotes ‘combined αCTLA-4 and αPD-1’, UC ‘ulcerative colitis’. **(D)**Relative fraction (range 0–1) of each cell type, stratified by tissue compartment, for all patients. Statistical testing for differences across groups separately per tissue compartment by Kruskal-Wallis test followed by Dunn’s post-hoc test with Benjamini-Hochberg false-discovery rate correction. Abbreviations cICI and UC as in (**C**).

We confirmed that all cell types were present across patients in comparable and biologically plausible frequencies (**Fig. 2C**). The ∼25% non-classified non-immune cells most likely represent stromal cells, which is numerically in line with previous IMC data in IBD patients.^24^ Based on epithelial masks, we additionally assigned each cell to the intra-epithelial or lamina propria compartment. As expected, we observed a high relative fraction of epithelial cells in epithelium and complete absence in lamina propria (**Fig. 2D**). Immune cells were relatively more abundant in lamina propria than epithelium, which was most evident for CD4^+^ T cells. CD8^+^ T cells were prevalent in both the lamina propria and intra-epithelial compartment (**Fig. 2D**). Specifically, CD8^+^ T cells dominated the intra-epithelial immune cell composition of αPD-1 and cICI-colitis and were significantly higher compared to αCTLA-4 colitis and UC (**Fig. 2D**). The same trend of CD8^+^ T cell dominance in αPD-1 and cICI-colitis could be observed in lamina propria. Epithelial macrophages were significantly reduced in cICI versus αCTLA-4 colitis, while relative abundance of CD4^+^ T cells, B cells, Tregs, dendritic cells and γδ T cells was similar between groups per compartment.

### Tissue resident memory CD8^+^ T cells are more abundant in epithelium than lamina propria and show high cytotoxic potential in most colitis types

The clinical course of cICI-colitis is often more acute and severe than ICI monotherapy-induced colitis.^1,10^ We explored if specific characteristics of the immune infiltrate could explain the clinical differences observed among colitis subtypes. Increased expression of cytotoxic effector protein granzyme B in CD8^+^ T cells has been reported in a broad range of inflammatory conditions, including Crohn’s disease and UC.^25^ In our study, CD8^+^ T cell cytotoxicity, quantified by CD8^+^ T cell granzyme B level, was significantly higher in cICI-than in ICI monotherapy-induced colitis or UC (**Fig. 3A,B**). The extent of CD8^+^ cytotoxicity did not correlate with any clinical or histological measure of colitis severity, including RHI, Common Terminology Criteria for Adverse Events (CTCAE) grade and Mayo endoscopic score (data not shown). We next evaluated the association between granzyme B levels in peripheral blood and the occurrence of irAEs, in particular ICI-colitis. Proteomic measurements in serum showed that irAEs after cICI indeed are associated with significantly higher granzyme A, B and H levels as compared to αPD-1 treatment alone (**Fig. 3C**). In addition, significantly higher circulating levels of IFN-γ, IL12Rβ1 and TNF-α were observed following cICI, compared to αPD-1 treatment in patients with irAEs. Although limited by smaller sample size, the same trend of an enhanced Th1-associated response with higher serum granzyme B levels was visible when only considering patients who developed ICI-colitis (**Supplementary Fig. 2**).

**Figure 3.**
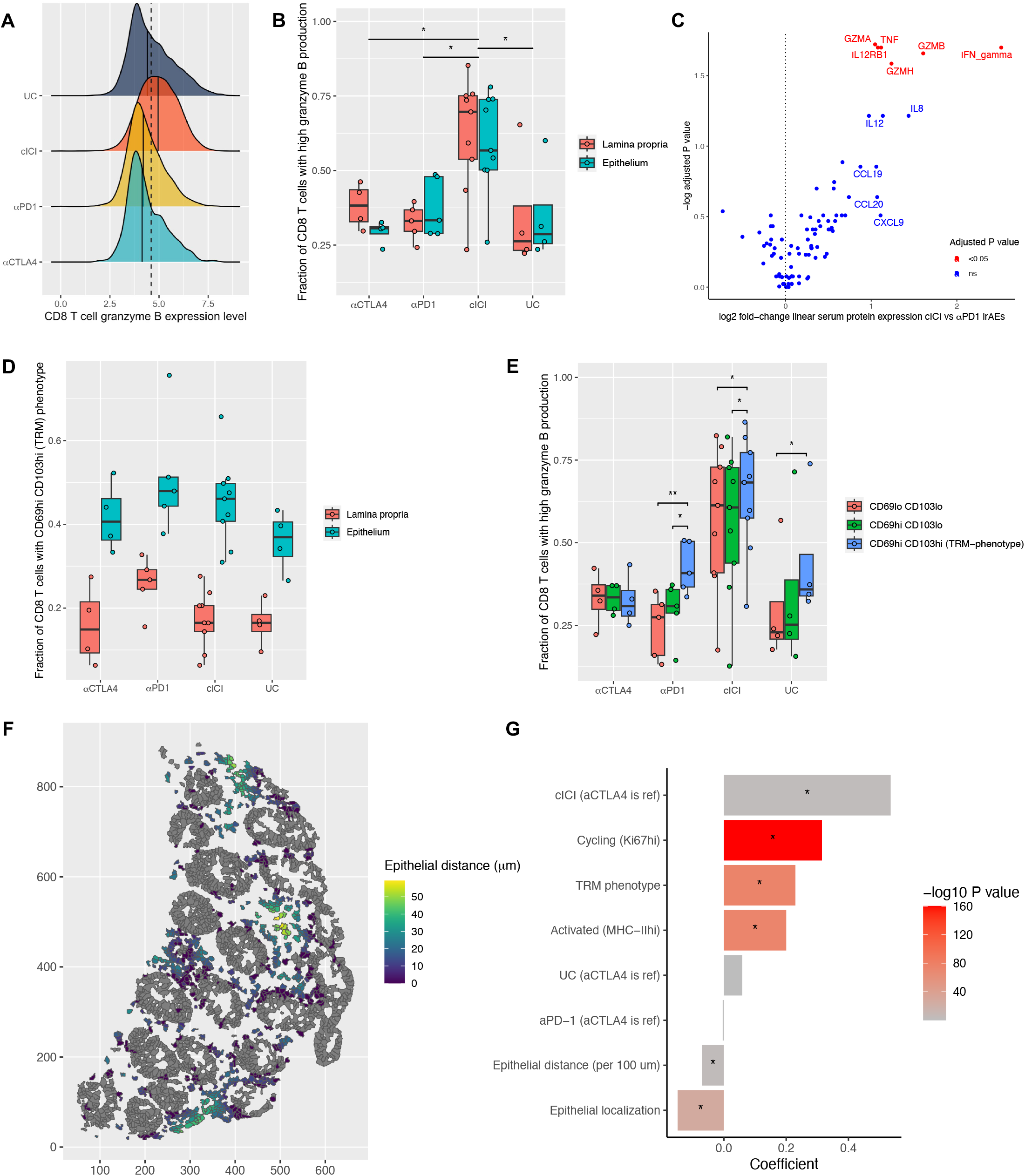
Phenotypical, functional and spatial CD8^+^ T cell characteristics related to cytotoxicity. **(A)**Density plot displaying granzyme B expression levels in CD8^+^ T cells, stratified by colitis subtype. Solid lines within each graph indicate the colitis subtype-specific median expression level. The dashed line displays the overall median expression level of all CD8^+^ T cells together (n=2,227–13,430 cells per colitis subtype). **(B)**Fraction of CD8^+^ T cells with higher-than-median cytosolic granzyme B levels across all CD8^+^ T cells, stratified by tissue compartment. Difference between groups (based on averaged lamina propria and epithelium data points) tested by one-way ANOVA, followed by Tukey’s post-hoc test. cICI denotes ‘combined αCTLA-4 and αPD-1’, UC ‘ulcerative colitis’, \**P*<0.05. **(C)**Volcano plot showing differential serum protein expression of 92 analytes in irAEs after combined αCTLA-4 and αPD-1 relative to αPD-1 monotherapy (only samples obtained upon irAE onset, n=39). Differential expression analyzed by Wilcoxon test with Benjamini-Hochberg false-discovery rate correction. **(D)**Fraction with T_RM_ phenotype of all CD8^+^ T cells, stratified by tissue compartment. No formal statistical comparisons were done. **(E)**Fraction of CD8^+^ T cells with higher-than-median cytosolic granzyme B levels across all CD8^+^ T cells, stratified by expression levels of CD69 and CD103. Pairwise comparisons were performed by paired Student’s t-tests with Benjamini-Hochberg false-discovery rate correction within each colitis subtype. cICI denotes ‘combined αCTLA-4 and αPD-1’, UC ‘ulcerative colitis’, \**P*<0.05, \*\**P*<0.01. **(F)**Representative example (ICI 7_A) of epithelial cells (grey) and CD8^+^ T cells in lamina propria, color-coded by distance to nearest epithelium (based on epithelial mask). **(G)**Visualization of coefficients and -log_10_(*P* values) of fixed effects. A mixed-effects model with fixed effects for all listed covariates and random intercepts for individual patients was developed to explain CD8^+^ T cell granzyme B production, using all 27,548 CD8^+^ T cells across n=22 patients. Coefficients >0.0 indicate that CD8^+^ granzyme B levels are higher in presence of (or for higher values of) the variable. \**P*<0.05.

Subsequently, we phenotypically characterized CD8^+^ T cells based on the expression of classical tissue residency markers CD69 and CD103.^26^ We grouped CD69^lo^CD103^lo^, CD69^hi^CD103^lo^ and CD69^hi^CD103^hi^ (termed T_RM_) CD8^+^ T cells.^27^ To this end, we defined “low expression” as below-median expression of CD69 or CD103 across all CD8^+^ T cells and *vice versa* for “high expression”. As expected, the classical T_RM_ phenotype was more abundant in epithelium than lamina propria in all colitis groups (**Fig. 3D**). Moreover, we found that granzyme B levels were highest in T_RM_ CD8^+^ T cells in all colitis subtypes, except αCTLA-4 colitis (**Fig. 3E**). Thus, our data suggest that the clinical disparities between different types of colitis may be partly due to difference in abundance of CD8^+^ T_RM_ cells and higher granzyme B levels in CD69^hi^CD103^hi^ T_RM_ than non-T_RM_ CD8^+^ T cells.

### Activated, cycling tissue resident memory CD8^+^ T cells below the epithelial border are key cytotoxic actors in colitis

IMC uniquely offers the opportunity to enrich phenotypical data with spatial information. We first assessed if CD8^+^ T cell tissue localization was associated with cytotoxic potential and therefore calculated distance to the nearest epithelial border for all CD8^+^ T cells, as shown by example in **Fig. 3F**. Then, we developed a mixed-effects model embedding clinical, phenotypical, spatial and functional (activation and proliferation status) covariates to delineate the contribution of each variable to total CD8^+^ granzyme B production. Based on this, an activated (MHC-II^hi^), cycling (Ki67^hi^) CD8^+^ T_RM_ phenotype was highly significantly (*P* < 10^−50^ for individual covariates) associated with higher granzyme B production (**Fig. 3G**). In adjusted analysis, cells adjacent to the epithelium produced more granzyme B than cells distant from epithelium and intra-epithelial cells (**Fig. 3G**), suggesting that inflammation was most pronounced at the lamina propria-epithelial interface. To evaluate how robust these findings were in relation to colitis subtype, we repeated the analysis stratified by colitis group. We found that the significant association between an activated, cycling CD8^+^ T_RM_ phenotype below the epithelial border and high granzyme B production was present across colitis subtypes, although the effect of mucosal localization was not significant in αPD-1 colitis (**Supplementary Fig. 3**). However, CD8^+^ T cell characteristics alone could not completely explain highest granzyme B production in cICI-colitis, as indicated by the effect through cICI treatment, adjusted for other covariates (**Fig. 3G**).

Based on increased serum levels of Th1-associated cytokines in cICI relative to αPD-1 irAEs (**Fig. 3C**), we hypothesized that enhanced CD4-help, T cell priming or activation might additionally underly high cICI-colitis cytotoxicity. Therefore, we also investigated if colitis subtypes differed with respect to cell-cell interactions using neighborhood analysis.^28^ We assessed interactions between all cell types and visualized interactions between epithelial cells, CD4^+^ and CD8^+^ T cells, CD4^+^ Tregs, dendritic cells (DCs), B cells and γδ T cells. General tissue organization in epithelium and lamina propria compartments was, as expected, reflected by significant avoidance between CD8^+^ T and epithelial cells, and CD4^+^ T and epithelial cells in both directions (**Supplementary Fig. 4**). Based on agglomerative clustering, patients could be divided in two groups. Significant interactions among adaptive immune cells, and DCs in addition influencing the adaptive response, presented as an important difference separating the group containing all UC patients from the other group (*P* = 0.03 by Fisher’s exact test, **Supplementary Fig. 4**). However, none of the ICI-colitis subtypes were significantly enriched in one of both adaptive immune cell interaction based subgroups. In summary, neighborhood analysis did not yield evidence in support of cICI-specific enhanced interactions between CD8^+^ T cells and other (adaptive) immune cells to additionally explain high cICI cytotoxicity.

**Figure 4.**
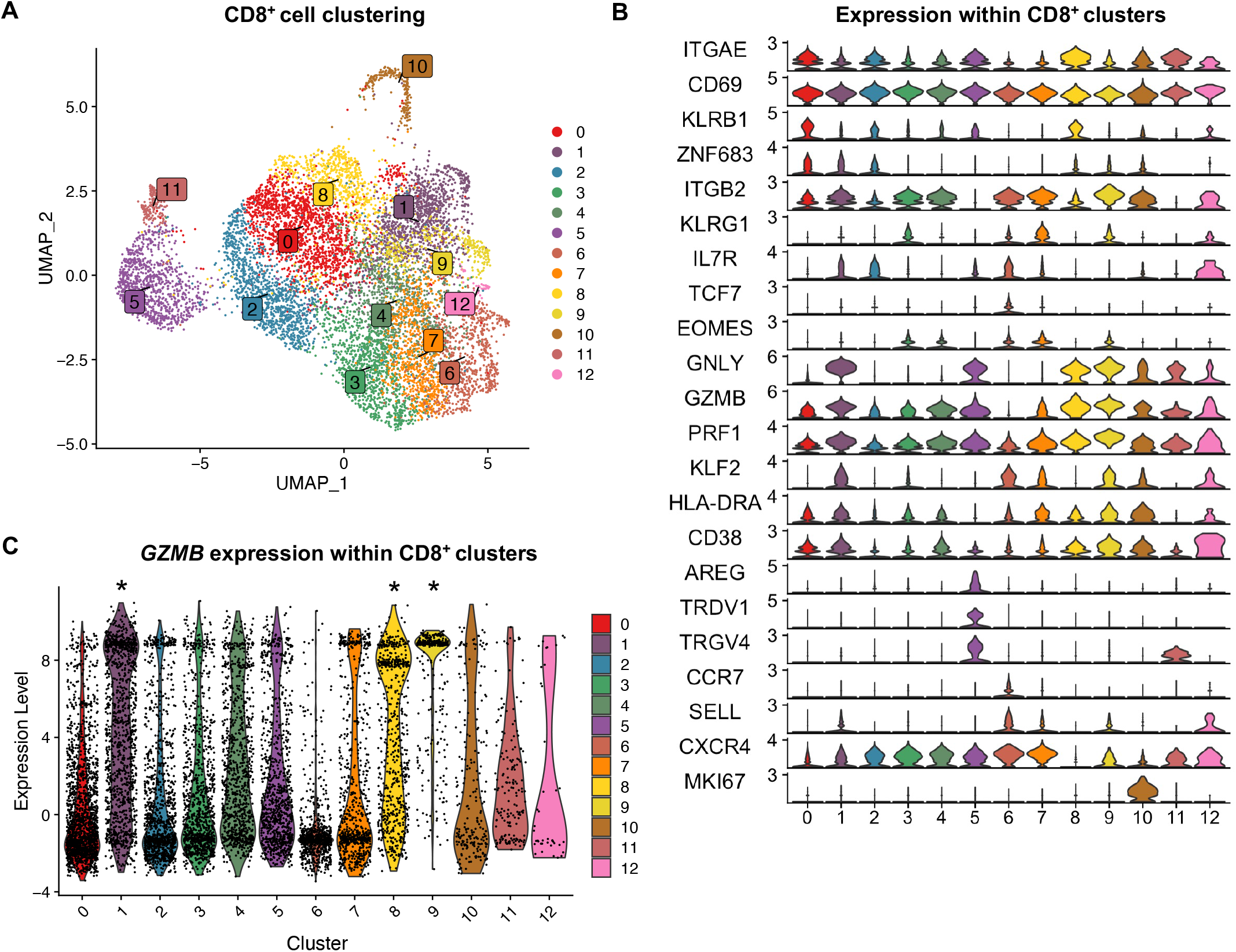
Reanalysis of CD8^+^ T cell clusters in colonic CD3^+^ single-cell RNA-sequencing data from ICI-colitis patients. **(A)**Uniform Manifold Approximation and Projection (UMAP) visualization showing sub-clustering of *CD8A*-expressing cells (*N*=8 ICI-colitis patients). **(B)**Stacked violin plot indicating expression of key subset-defining genes within *CD8A*-expressing clusters (*N*=8 ICI-colitis patients). **(C)**Violin plot indicating *GZMB* expression in CD8^+^ cell clusters; clusters with statistically significantly higher *GZMB* expression compared to all other CD8^+^ cell clusters together are indicated with an asterisk. Adjusted *P* values are 3e-137, 5e-105 and 5e-82 for respectively cluster 1, 9 and 8, by Wilcoxon test with Benjamini-Hochberg false-discovery rate correction (*N*=8 ICI-colitis patients).

### Activated CD8^+^ T_(RM)_ cells are confirmed as drivers of ICI-colitis at the transcriptomic level

To validate CD8^+^ T_RM_ cells as important mediators of inflammation in ICI-colitis, we reanalyzed colonic CD3^+^ scRNA-seq data from eight patients with ICI-colitis after αCTLA-4 with or without αPD-1 therapy.^14^ We selected *CD8A*-expressing cells from the CD3^+^ T cell pool, clustered the CD8^+^ T cells and annotated clusters based on the top 6 cluster-defining genes along with other known markers (**Fig. 4A,B, Supplementary Fig. 5**). Clusters 0 and 8 represented CD103^+^ T_RM_ clusters (*ITGAE, KLRB1*), while clusters 3 and 7 were identified as CD103^-^ T_RM_ subsets (*ITGB2, KLRG1*). Other subsets included CD8^+^ γδ T cells (cluster 5; *AREG, TRDV1, TRGV4*), central memory CD8^+^ T cells (cluster 6; *CCR7, SELL, CXCR4*), a cycling subset (cluster 10; *MKI67*) and highly cytotoxic effector T cells (clusters 1 and 9; *GZMB, PRF1*) with cluster 9 being terminally differentiated (*KLRG1*^*+*^, *IL7R*^*-*^). *GZMB* expression was significantly higher in clusters 1, 8 and 9 compared to all other clusters (**Fig. 4C**), importantly including a cluster representing activated CD103^+^ T_RM_ cells (cluster 8; *CD38, ITGAE, KLRB1*). Consistent with IMC data, the CD103^-^ T_RM_ subsets (clusters 3 and 7) featured low *GZMB* expression (**Fig. 4C**). Our sub-clustering results fit the possible differentiation path from T_RM_ to cytotoxic effector cells as discovered by Luoma et al.^14^ An important role for activated CD103^+^ CD8^+^ T_RM_ cells in granzyme B mediated cytotoxicity is thus confirmed at the transcriptomic level.

## Discussion

ICI-colitis and UC show several clinical and histological similarities, but it is unknown to what extent they share a common pathophysiology. Here, we described our approach combining DAPI imaging and IMC to acquire high-resolution, highly multiplexed, single-cell images of inflamed colon from patients with αCTLA-4-, αPD-1-, cICI- or ulcerative colitis. We explored on the protein level what cell types are involved in those subtypes of colitis and assessed phenotypical, spatial and functional T cell characteristics.

In line with other studies, we found that CD8^+^ T cells are relatively increased in αPD-1 and cICI-colitis compared to UC.^15,29^ We confirmed that activated cycling CD8^+^ T_RM_ cells are important drivers of inflammation in colitis and additionally our data suggest that their cytotoxic potential was highest below the epithelial border. cICI-colitis exhibited higher tissue CD8^+^ T cell granzyme B levels compared to all other colitis types, which was corroborated in serum of cICI- and αPD-1-treated patients who developed irAEs including ICI-colitis. The fact that clinical and histological colitis severity indices and CD8^+^ T cell cytotoxicity were not correlated, may suggest that CD8^+^ granzyme B levels possibly reflect the induction of inflammation, while factors beyond CD8^+^ T cell biology may collectively determine clinical severity and thus the need for escalated immunosuppressive therapy. Besides, nor CD8^+^ T cell characteristics, nor cell-cell interactions could completely account for the colitis subtype-related differences in cytotoxicity. Nevertheless, others have similarly found enhanced CD8^+^ T cell *GZMB* or *GZMK* expression in cICI-colitis relative to αPD-1 colitis.^15,16^ We previously showed that, in contrast to αPD-1, irAEs after cICI are strongly associated with peripheral blood effector memory CD4^+^ T cell proliferation amidst a mainly Th1-associated response.^30^ In the present study, we confirmed that cICI irAEs come with stronger peripheral Th1-skewing than αPD-1 irAEs. This indicates that enhanced cytotoxicity observed in cICI-colitis might be the result of reinvigorated CD4^+^ T cell help, potentially lowering the threshold for CD8^+^ T cell activation. Although our study revealed no clues to support this hypothesis, higher *CD28* and *TNFRSF4* (encoding CD134) expression in CD8^+^ T cells of cICI-colitis than αPD-1 colitis shown previously, suggests enhanced T cell receptor signaling in cICI-colitis.^16^ Of note, whether CD8^+^ T_RM_ cells may have been activated elsewhere before they entered the tissue, or CD8^+^ T_RM_ cells are directly targeted via PD-1 by ICI therapy, or both, cannot be concluded from these studies.

We detected higher CD8^+^ T cell cytotoxicity at the lamina propria-epithelial interface. Since intraepithelial cells have been shown to migrate dynamically between epithelium and lamina propria,^31^ our observation is compatible with the finding that especially intraepithelial CD103^+^ CD8^+^ T cells adopt a cytotoxic profile in Crohn’s ileitis,^21^ and that the highest expression of UC-associated loci is found in intraepithelial CD8+ T cells of UC patients.^32^ Although data are conflicting, others have shown substantial clonal overlap and transcriptional similarity between lamina propria and intra-epithelial lymphocytes in the mucosa of healthy controls.^33,34^ Distinguishing two “canonical” T_RM_ populations classified as CD103^-^ (*KLRG1, ITGB2*) and CD103^+^ (*ITGAE, KLRB1*) T_RM_ subsets, in healthy tissue, the former were associated with highest cytotoxic potential, while the latter produced multiple cytokines like TNF-α and IFN-γ simultaneously.^33^ Interestingly, we showed increased granzyme B production in CD8^+^ T cells with *higher* CD103 expression. Our findings are corroborated by others who reported greatest increase in expression of *GZMB* and *IFNG*, or *IL17A* and *IL26* in *ITGAE*-expressing subsets in ICI-colitis.^15,16^ This may present as a dissimilarity with Crohn’s disease, in which data are conflicting as to whether CD103^+^ or CD103^-^ CD8^+^ T_RM_ cells are most cytotoxic.^21,35,36^ Although some studies reported decreased abundance of intestinal CD8^+^ T_RM_ cells in patients with ICI-colitis compared to ICI-treated patients without colitis,^14,16^ cytotoxic lymphocytes newly emerging in tissue were clonally related to *bona fide* T_RM_ cells.^14^ Compared to active inflammation in UC, CD8^+^ T_(RM)_ cells are more abundant and more activated in ICI-colitis.^15,29^ In summary, an accumulating body of evidence points towards CD8^+^ T_RM_ cells as drivers of inflammation, potentially playing an even more prominent role in ICI-colitis than UC.

Thus, intestinal CD103^+^ CD8^+^ T_RM_ cells might serve as a therapeutic target in UC and ICI-colitis. Blocking the integrin pair α4β7 with vedolizumab has proven effective in treating both UC and ICI-colitis,^6,37^ but does not target αE^+^ (CD103^+^) CD8^+^ T_RM_ cells. Etrolizumab is a monoclonal antibody against the β7 integrin subunit, with activity against α4β7^+^ and αEβ7^+^ cells. Based on cell type signature scores applied to bulk RNA-seq data from IBD patients, etrolizumab reduced the number of both intestinal CD103^-^ and CD103^+^ CD8^+^ T cells.^36^ However, to date clinical results with etrolizumab in Crohn’s and ulcerative colitis are disappointing and it is currently unclear if the drug will be clinically taken any further.^38,39^ A possible explanation for the lack of efficacy might be that with the depletion of intestinal CD103^+^ T_RM_ cells, not only pro-inflammatory but also homeostasis-promoting T_RM_ subsets are lost. Apart from the limited efficacy of β7 integrin blockade in IBD, this strategy may carry a risk in itself when used in patients with cancer. The presence of CD103^+^ tumor-infiltrating lymphocytes (TILs) have been associated with better outcomes in most cancer types, especially of epithelial origin.^40^ TILs associated with tumors of other origin, including melanoma, also express *ITGAE* and *ITGB7* which can together form dimerized CD103.^41,42^ Directly targeting αEβ7 could negatively impact intra-tumoral retention of TILs and thus compromise anti-tumor immunity. In order to avoid this kind of adverse effects of T_RM_-directed treatment, future research should focus on phenotypical differences between irAE-tissue and tumor-associated T_RM_ cells.

As ICI-colitis appears dominantly Th1/Tc1-mediated disease,^14,15,17,43^ Jak inhibition could be another treatment strategy in ICI-colitis.^44^ Its safety from a tumor response perspective is presently not well established, however. Finally, upregulation of *IL17* and *IL26* differentiated cICI from αPD-1 colitis.^16^ Involvement of the IL-23/IL-17 axis has been demonstrated in various irAEs and is more pronounced after cICI.^30,45,46^ Paradoxically, direct IL-17 blocking, e.g. with secukinumab, has been shown to induce intestinal inflammation.^47^ Pro-homeostatic IL-17 production is independent of IL-23,^48^ and therefore more upstream interventions directed against IL-23, such as ustekinumab, or the IL-6 receptor inhibitor tocilizumab might hold promise.

In conclusion, our study importantly contributes to our understanding of the pathophysiology underlying ICI-colitis and UC, including disparities among colitis subtypes, through integrated analysis of phenotypical, functional and spatial aspects. In this way, we underpin the role of CD8^+^ T_RM_ cells as potentially targetable drivers of ICI-colitis.

### Limitations of the study

Our study has two main limitations. First, this study did not include healthy controls. However, we confirmed antibody binding specificity both with immunofluorescence on control tissue and after isotope labeling with IMC using a tissue microarray, including uninflamed colon tissue, that was included on every slide with study participant tissue. Moreover, studies comparing colon mucosa from healthy controls with mucosa from ICI-colitis cases found CD8^+^ T_(RM)_ cell numbers to be comparable or higher in colitis. In addition, *GZMB* expression in different CD8^+^ subsets appears to be considerably higher in colitis.^14,15,29^ Second, our IMC antibody panel lacked stromal markers. For this reason, cell types such as fibroblasts could not be classified. However, the primary goal of this study was to compare different types of ICI-colitis and UC, delineate the abundance of different immune cells for those colitis subtypes and relate immune cell phenotypes to their cytotoxic potential *within* patients. Moreover, our lineage definition approach accounted for lacking markers of non-immune cells and, therefore, mis-classification of key immune cells of interest such as CD8^+^ T cells could be minimized.

## Data Availability

An R markdown file with code used for IMC data processing and analysis has been deposited on Zenodo, publicly accessible via DOI: 10.5281/zenodo.7858216. In compliance with national legislation, upon acceptance of the manuscript data produced in the present study will become available online (DataverseNL) with restrictions.

## Author contributions

Conceptualization: ME, EB, JL, BO, YV, KS, FW Methodology: ME, MB, JD Formal analysis and visualization: ME Biobank patient inclusions and sampling: RV, ME Performing experiments: JL, MA, ED Histopathologic scoring/annotation: ML Writing original draft: ME Reviewing & editing: all authors Approval of final manuscript: all authors Supervision: KS, FW Funding acquisition: EB, BO, YV, FW

## Acknowledgements

We would like to thank Domenico Castigliego for the preparation of colon tissue slides.

## Declaration of interests

EB: received research funding from Pfizer and is supported by the Alexandre Suerman stipend for MD/PhD candidates of the UMC Utrecht.

YV: has received speaker fees from Johnson & Johnson, research funding from Galapagos and a Public Private Partnership grant from Health Holland (#TKI2017), with TigaTx B.V.

KS: has advisory relationships with Bristol Myers Squibb, Novartis, MSD, Pierre Fabre, AbbVie and received honoraria from Novartis, MSD and Roche. Research funding: BMS, Philips, TigaTx. All paid to institution.

FW: has received advisory/speaker fees from Takeda, and Johnson&Johnson, and has received research funding from BMS, Takeda, Sanofi, Pfizer, Galapagos, and Leo Pharma.

No potential conflicts of interest were disclosed by the other authors.

## Funding

This work received funding from the Dutch Society of Gastroenterology (NVGE; Gastrostart grant) and NWO Gravitation 0.24.001.028, cancergenomicscenter.nl (to YV).

## Methods

### Code availability

An R markdown file with code used for IMC data processing and analysis has been deposited on Zenodo, publicly accessible via DOI: 10.5281/zenodo.7858216.

### Study participants

Colon biopsies were obtained from patients treated with αCTLA-4 monotherapy, αPD-1 monotherapy or combined αCTLA-4 and αPD-1 (cICI) who developed ICI-colitis. All biopsies were obtained during routine clinical procedures. Colon biopsies from UC patients and corresponding clinical data were obtained through the University Medical Center Utrecht Research Data Platform. In suspected ICI-colitis patients, biopsies were taken only from inflamed mucosal sites (or at-random in case of endoscopically uninflamed mucosa) in the left-sided colon. Mayo endoscopic scores were retrieved from endoscopy reports or retrospectively assessed from endoscopic images.^49^ Tissue samples from ICI-colitis patients were only used if histological assessment confirmed active inflammation and no concurrent gastro-intestinal infection was suspected. UC patients from whom samples were included had to be steroid- and biological-free in the six months prior to endoscopy. Serum was obtained from patients participating in the UNICIT biobank study.^30^ Serum samples collected at baseline and ±6 weeks into treatment (for irAE-free patients) or upon development of Common Terminology Criteria for Adverse Events (CTCAE) v5^50^ grade ≥2 irAEs demanding hospitalization or ≥0.5 mg/kg steroids were selected for multiplex immunoassay. All patients from whom serum is used participated in the UNICIT biobank study and provided written informed consent in accordance with the Declaration of Helsinki. Ethical approval was received from the UMC Utrecht Biobank Review Committee (*Toetsingscommissie Biobanken* [TC-bio] 18-123) and permission to use human specimens form this biobank was granted (TC-bio 19-704). Use of anonymous or coded leftover material for scientific purposes is part of the standard treatment contract on an opt-out basis with patients in our hospital. Approval for use of clinical metadata from the pseudonymized UC patients was obtained through TC-bio 18-676.

### Sample collection and histological assessment

For each participant, two adjacent 4 μm-thick tissue sections were prepared from colon biopsies fixed in 10% formalin and paraffin-embedded (FFPE). One slide was stained with hematoxylin & eosin (H&E) while the other was used for nuclear imaging and IMC. Serum samples were isolated and stored at -80°C within 4 hours after blood collection. H&E-stained slides of all included samples were assessed by an experienced gastro-intestinal pathologist (M.L), who annotated lymphoid follicles in the regions used for IMC and assessed disease activity employing the Geboes scores,^51^ validated for UC but not for ICI-colitis.^52^ These scores were then used to calculate the Robarts Histopathology Index (RHI).^53^ Histological scoring was based on the highest inflammation score found within the entire tissue section available. Hence, IMC regions did not necessarily feature all histologic characteristics that prompted a certain Geboes score in the same patient.

### Slide preparation for microscopy and IMC

After confirming antibody binding specificity with immunofluorescence on tonsil and colon FFPE sections, antibodies were conjugated to lanthanide isotopes with the MaxPar antibody labeling kit (Fluidigm, San Francisco, CA, USA) according to the manufacturer’s protocols. Tissue microarrays comprising different types of tissue, including non-inflamed colon, ovarian cancer and tonsil, were present on the FFPE slides used for IMC to validate antibody binding and specificity. FFPE tissue slides were baked (1.5 hours, 60°C), deparaffinized with xylene (20 min.) and rehydrated in a gradient of ethanol (100%; 10 min., followed by 95%, 80%, 70%; 5 min. each). Slides were then washed in MilliQ water (3 min.) and phosphate-buffered saline (PBS) containing 0.1% Tween (PBST; 10 min.). Heat-induced epitope retrieval in 10mM Tris with 1mM EDTA (pH 9.5, 30 min., in a 95°C water bath), cooling to 70°C and washing in PBST (10 min.) were followed by blocking with 3% bovine serum albumin (BSA) and 1:100 Human TruStain FcX in PBST (1 hour, room temperature [RT]). After removal of the blocking buffer, slides were incubated overnight with the antibody cocktail (**Supplementary Table 2**) in PBST with 0.5% BSA (4°C). Slides were then washed three times in PBST. Next, slides were stained with 1:400 DNA-intercalator (Ir-191/Ir-193) and 1:1000 4′,6-diamidino-2-phenylindole (DAPI, 60 min., RT) in PBS, followed by two washes with MilliQ water, and air-dried.

### Fluorescent microscopy imaging

Slides were imaged on a Zeiss Z1 imager using a 20x dry objective (Zeiss, EC Plan-NEOFLUAR 0.5 NA, 420350-9900) with mercury lamp as light source, a 49 filter set and an Axiocam 503 mono camera system. Using ZEN software (2.6), images were acquired in a tiled Z-stack format (9 Z-slices) with 10% overlap between tiles and exported to individual 16-bit tiff tiles. Single in-focus images were created with the Extended Depth of Field plugin in Fiji and tile images were stitched using the MIST algorithm in Fiji.^54–56^

### Imaging mass cytometry and cell segmentation

After acquisition of DAPI images, slides were rinsed in MilliQ water and counterstained with 0.1% toluidine blue (5 min., RT), washed with MilliQ water and air-dried. Mass cytometry of ∼1 mm^2^ regions per patient was performed on a Helios (Fluidigm) mass cytometer connected to a Hyperion (Fluidigm) laser ablation module (ablation frequency 200 Hz). Imctools was used to convert data to 32-bit tiff files. For two control tissues included within the tissue microarray, single channel expression data are shown in **Supplementary Fig. 6A**,**B**. Single cell segmentation was performed using the MATISSE segmentation pipeline, combining IMC and DAPI images, as described elsewhere.^23^ This combined approach achieves superior cell segmentation compared to IMC-based segmentation alone.^22^ Briefly, high-resolution DAPI images were registered to IMC images. Training data for IMC images (with annotations for epithelial and non-epithelial cell membranes, epithelial and non-epithelial nuclei and background) and DAPI images (only nuclear annotations) were created by two researchers (M.E. and J.L.). IMC channels used for annotations were E-cadherin, CD68, CD14, CD20, CD45, CD45RA, CD45RO, CD4, CD8α, IL-17A, CD3, CD69 and Ir-193. Probability maps for nuclear and cellular objects were created in Ilastik,^57^ and cell segmentation was performed with CellProfiler using these probability maps.^58^ Next, based on all pixels within a segmented cell, mean IMC signal expression for all channels was extracted for all segmented cells with R.

### IMC data clean-up and normalization

A step-by-step description of our approach for data clean-up, normalization, scaling and annotation is provided in the R markdown document uploaded at Zenodo (DOI: 10.5281/zenodo.7858216). First, artefacts (e.g., antibody aggregates), lymphoid follicles as indicated by the gastro-intestinal pathologist and submucosa were manually annotated in Fiji by two researchers (M.E. and J.L.). Coordinates for boundaries of epithelial regions were extracted from Fiji using the E-cadherin channel, smoothened with Gaussian blur (σ = 2 pixels [μm]). Next steps were exclusively performed in R. Data were natural-log transformed. Single cells within artefact, lymphoid follicle or submucosa tissue regions were excluded and remaining events were labeled as either “intra-epithelial” or “lamina propria” based on their intersection with epithelial masks using the *sf* package.^59^

### IMC data scaling and cell lineage annotation

For data intensity scaling, we calculated scaling factors, derived from modal marker intensities, for each channel and each patient separately, since we assumed that channel intensity variability resulted 2from both patient- and channel-specific sources. Then, all data were linearly scaled, effectively aligning channel-specific single-cell expression distributions among patients. Normalization and scaling results were visually checked by histogram representations of single-cell data for all patients, as shown in **Supplementary Fig. 7A**. We confirmed that scaling factors represented a normal distribution, both over channels and over patients (**Supplementary Fig. 7B**). We also verified that markers for which expression correlated within cells were also biologically related (**Supplementary Fig. 7C**).

Subsequently, channel positivity thresholds were defined for markers that were selected for lineage determination. For markers with bimodal expression, a threshold separating the marker-positive and -negative populations was applied. For unimodally distributed markers in which “true positive signal” is contained in the heavy right tail, a normal distribution was derived from the left half of the histogram using the “full width at half maximum” method. Then, a threshold for marker positivity was conservatively set at +1.5 standard deviation (SD). Next, candidate cell type annotations were generated using the Boolean rules in the Table below. Due to imaging resolution, single-cell signal will inevitably contain signal of neighboring/overlapping cells, complicating unsupervised cell type annotation based mostly on membrane markers, especially in dense areas. Therefore, we developed a supervised approach in which each cell could be assigned multiple different candidate cell types. Next, expression levels of key markers (indicated in bold in the Table below) were ranked across all cells from all patients pooled together. For each individual cell, ranks of key markers were then compared between the candidate cell types. The key marker ranked highest eventually determined the assigned cell type label for each single cell. Cells that did not fit the criteria for any of the cell types were labeled “non-classified”, or “other immune cell” if they expressed any of CD45 isoforms.

### Boolean rules for candidate cell type annotation

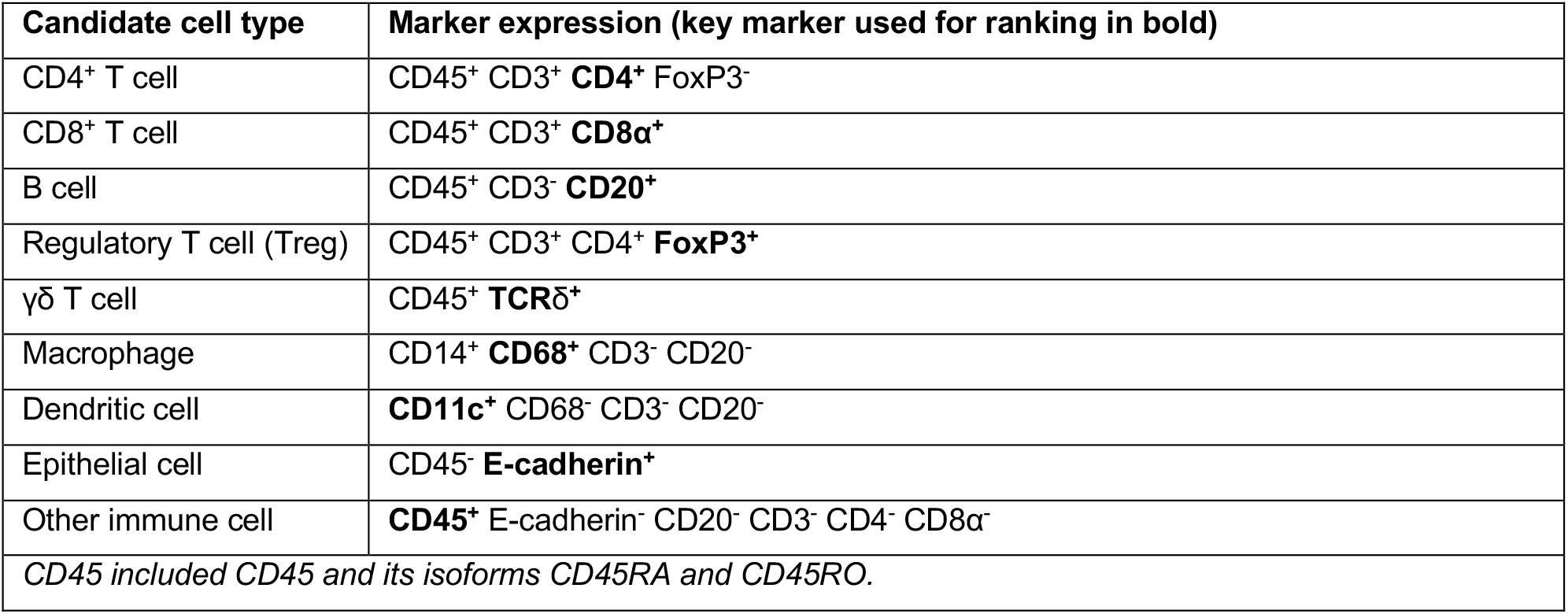

After cell type annotation, we excluded one UC patient (IBD 1_B) from further analysis because of low signal-to-noise ratio and large within-image intensity variation across channels, which yielded ill-annotated cells (**Supplementary Fig. 8**).

### IMC data dimensional reduction

Uniform Manifold Approximation and Projection (UMAP) visualizations were created with *umap()* and 15 neighbors for a randomly drawn subset containing 10% of all cells.^60^ Mean single-cell intensities of all channels, except RORγt was used, because data normalization was imperfect for this marker, artificially leading to patient-based separation.

### IMC neighborhood analysis

Cell-cell interactions for all cell types (including non-classified cells) were investigated by neighborhood analysis as described in Schapiro et al. using the *neighbouRhood* package with n=10,000 permutations.^28^ An agglomerative clustering heatmap was built based on interactions of prespecified relevant cell types. Hierarchical clustering was performed using Ward’s method based on Manhattan distance.

### Multiplexed proteomics

Normalized protein expression (NPX) levels of 92 cytokines and chemokines was measured in serum by proximity extension immunoassay using the Olink^®^ Target 96 Immuno-Oncology panel.^61^ This technique is based on the ligation of two complementary DNA strands that are coupled to panel-specific antibodies. After ligation, double-stranded DNA is polymerase chain reaction (PCR) amplified and the detected DNA tags are following corrections for extension- and interpolation controls translated to NPX values on an (inherently) relative log_2_-scale. Thus, 1 NPX difference between two samples represents a doubling of protein expression between samples. To correct for batch effects across plates, data were normalized using 12 bridging controls that were included on all measured Olink plates. NPX values below the limit-of-detection (LOD) were replaced with 0.5×LOD.

### scRNA-seq data reanalysis

Previously published (GSE144469)^14^ CD3^+^ T cell scRNA-seq data from 8 αCTLA-4 monotherapy or combined/sequentially ICI-treated patients who developed ICI-colitis was reanalyzed using *Seurat* v4.^62^ Cells with 200-3,000 features and <10% mitochondrial transcripts were kept for further analysis. SCTransform() was used on each sample individually with vars.to.regress=‘percent.mt’ and method=‘glmGamPoi’ to correct for technical variation including batch effect.^63^ Then all objects were integrated using SelectIntegrationFeatures() with 2,000 variable features and clustering was performed with FindNeighbors() to construct a K-nearest neighbor (KNN) graph using the first 30 principal components, followed by FindClusters() with resolution=0.8. CD8^+^ T cell clusters were selected based on mean-normalized *CD4* expression <1 and *CD8A* expression >1, with expression >1 indicating above-average expression. Then we performed clustering within the CD8^+^ cell pool using the same parameters. Cell clusters were annotated based on the top 6 markers defining each cluster, using FindAllMarkers() with min.pct=0.1 and logfcthreshold=0.01, along with known population-defining markers.

### Statistical analysis

All analyses were performed in R version 4.2.0. Continuous variables were compared between multiple groups with a one-way ANOVA followed by Tukey’s post-hoc test (for normally distributed data) or the Kruskal-Wallis test followed by Dunn’s post-hoc test with Benjamini-Hochberg false-discovery rate (FDR) correction (for non-normally distributed data). Spearman’s rank coefficient was used for correlations between two continuous variables and associations between categorical variables were tested with Fisher’s exact test. Differences in normalized protein expression (NPX) obtained in the serum proteomics measurements was compared by Wilcoxon tests with Benjamini-Hochberg FDR correction. The *nlme* package (v3.1-158) was used to analyze CD8^+^ granzyme B production by a linear mixed-effects model with random intercept for patients and fixed effects for all covariates, fit by restricted maximum likelihood. Two-sided *P*<0.05 was considered statistically significant. Statistical details of individual analyses are reported in the figure legends. Unless otherwise specified, n represents the number of participants.

## Notes

### Author Declarations

All patients from whom serum is used participated in the UNICIT biobank study and provided written informed consent in accordance with the Declaration of Helsinki. The Biobank Review Committee (Toetsingscommissie Biobanken [TC-bio]) of University Medical Center Utrecht gave ethical approval for this work (TC-bio 18-123) and granted permission to use human specimens form this biobank (TC-bio 19-704). Use of anonymous or coded leftover material for scientific purposes is part of the standard treatment contract on an opt-out basis with patients in our hospital. Approval for use of clinical metadata from the pseudonymized UC patients was obtained through TC-bio 18-676.

## References

1. Chan KK, Bass AR. Autoimmune complications of immunotherapy: pathophysiology and management. The BMJ. 2020;369:m736. doi:10.1136/bmj.m736

2. Arnaud-Coffin P, Maillet D, Gan HK, et al. A systematic review of adverse events in randomized trials assessing immune checkpoint inhibitors. Int J Cancer. 2019;145(3):639–648. doi:10.1002/ijc.32132

3. Wang DY, Salem JE, Cohen J v., et al. Fatal Toxic Effects Associated With Immune Checkpoint Inhibitors. JAMA Oncol. 2018;4(12):1721. doi:10.1001/jamaoncol.2018.3923

4. Schneider BJ, Naidoo J, Santomasso BD, et al. Management of Immune-Related Adverse Events in Patients Treated With Immune Checkpoint Inhibitor Therapy: ASCO Guideline Update. Journal of Clinical Oncology. 2021;39(36):4073–4126. doi:10.1200/JCO.21.01440

5. Haanen J, Obeid M, Spain L, et al. Management of toxicities from immunotherapy: ESMO Clinical Practice Guideline for diagnosis, treatment and follow-up. Annals of Oncology. 2022;33(12):1217–1238. doi:10.1016/j.annonc.2022.10.001

6. Zou F, Faleck D, Thomas A, et al. Efficacy and safety of vedolizumab and infliximab treatment for immune-mediated diarrhea and colitis in patients with cancer: a two-center observational study. J Immunother Cancer. 2021;9(11):e003277. doi:10.1136/jitc-2021-003277

7. Yu B, Zhao L, Jin S, He H, Zhang J, Wang X. Model-Based Meta-Analysis on the Efficacy of Biologics and Small Targeted Molecules for Crohn’s Disease. Front Immunol. 2022;13. doi:10.3389/fimmu.2022.828219

8. Vasudevan A, Gibson PR, Langenberg DR van. Time to clinical response and remission for therapeutics in inflammatory bowel diseases: What should the clinician expect, what should patients be told? World J Gastroenterol. 2017;23(35):6385–6402. doi:10.3748/wjg.v23.i35.6385

9. Lo Y chun, Price C, Blenman K, Patil P, Zhang X, Robert ME. Checkpoint Inhibitor Colitis Shows Drug-Specific Differences in Immune Cell Reaction That Overlap With Inflammatory Bowel Disease and Predict Response to Colitis Therapy. Am J Clin Pathol. Published online 2021:Epub ahead of print. doi:10.1093/ajcp/aqaa217

10. Wang DY, Mooradian MJ, Kim D, et al. Clinical characterization of colitis arising from anti-PD-1 based therapy. Oncoimmunology. 2019;8(1):e1524695. doi:10.1080/2162402X.2018.1524695

11. Ibraheim H, Baillie S, Samaan MA, et al. Systematic review with meta-analysis: effectiveness of anti-inflammatory therapy in immune checkpoint inhibitor-induced enterocolitis. Aliment Pharmacol Ther. 2020;52(9):1432–1452. doi:10.1111/apt.15998

12. Bamias G, Delladetsima I, Perdiki M, et al. Immunological Characteristics of Colitis Associated with Anti-CTLA-4 Antibody Therapy. Cancer Invest. 2017;35(7):443–455. doi:10.1080/07357907.2017.1324032

13. Coutzac C, Adam J, Soularue E, et al. Colon immune-related adverse events: Anti-CTLA-4 and anti-PD-1 blockade induce distinct immunopathological entities. J Crohns Colitis. 2017;11(10):1238–1246. doi:10.1093/ecco-jcc/jjx081

14. Luoma AM, Suo S, Williams HL, et al. Molecular Pathways of Colon Inflammation Induced by Cancer Immunotherapy. Cell. 2020;182(3):655-671.e22. doi:10.1016/j.cell.2020.06.001

15. Sasson SC, Slevin SM, Cheung VTF, et al. IFNγ-producing CD8+ tissue resident memory T cells are a targetable hallmark of immune checkpoint inhibitor-colitis. Gastroenterology. Published online 2021:Epub ahead of print.

16. Fisher Thomas M, Slowikowski K, Manakongtreecheep K, et al. Altered interactions between circulating and tissue-resident CD8 T cells with the colonic mucosa define colitis associated with immune checkpoint inhibitors. bioRxiv. Published online September 20, 2021. doi:https://doi.org/10.1101/2021.09.17.460868

17. Nahar KJ, Marsh-Wakefield F, Rawson R v., et al. Distinct pretreatment innate immune landscape and posttreatment T cell responses underlie immunotherapy-induced colitis. JCI Insight. 2022;7(21). doi:10.1172/jci.insight.157839

18. Knauss A, Gabel M, Neurath MF, Weigmann B. The Memory T Cell “Communication Web” in Context with Gastrointestinal Disorders—How Memory T Cells Affect Their Surroundings and How They Are Influenced by It. Cells. 2022;11(18):2780. doi:10.3390/cells11182780

19. Noble A, Durant L, Hoyles L, et al. Deficient Resident Memory T Cell and CD8 T Cell Response to Commensals in Inflammatory Bowel Disease. J Crohns Colitis. 2020;14(4):525–537. doi:10.1093/ecco-jcc/jjz175

20. Roosenboom B, Wahab PJ, Smids C, et al. Intestinal CD103+CD4+ and CD103+CD8+ T-Cell Subsets in the Gut of Inflammatory Bowel Disease Patients at Diagnosis and During Follow-up. Inflamm Bowel Dis. 2019;25(9):1497–1509. doi:10.1093/ibd/izz049

21. Lutter L, Roosenboom B, Brand EC, et al. Homeostatic Function and Inflammatory Activation of Ileal CD8+ Tissue-Resident T Cells Is Dependent on Mucosal Location. Cell Mol Gastroenterol Hepatol. 2021;12(5):1567–1581. doi:10.1016/j.jcmgh.2021.06.022

22. Baars MJD, Sinha N, Amini M, et al. MATISSE: a method for improved single cell segmentation in imaging mass cytometry. BMC Biol. 2021;19:99.

23. Krijgsman D, Sinha N, Baars MJD, van Dam S, Amini M, Vercoulen Y. MATISSE: An analysis protocol for combining imaging mass cytometry with fluorescence microscopy to generate single-cell data. STAR Protoc. 2022;3(1):101034. doi:10.1016/j.xpro.2021.101034

24. Kondo A, Ma S, Lee MYY, et al. Highly Multiplexed Image Analysis of Intestinal Tissue Sections in Patients With Inflammatory Bowel Disease. Gastroenterology. 2021;161(6):1940–1952. doi:10.1053/j.gastro.2021.08.055

25. Boivin WA, Cooper DM, Hiebert PR, Granville DJ. Intracellular versus extracellular granzyme B in immunity and disease: challenging the dogma. Laboratory Investigation. 2009;89(11):1195–1220. doi:10.1038/labinvest.2009.91

26. Lutter L, Hoytema van Konijnenburg DP, Brand EC, Oldenburg B, van Wijk F. The elusive case of human intraepithelial T cells in gut homeostasis and inflammation. Nat Rev Gastroenterol Hepatol. 2018;15(10):637–649. doi:10.1038/s41575-018-0039-0

27. Kumar B V, Ma W, Miron M, et al. Human Tissue-Resident Memory T Cells Are Defined by Core Transcriptional and Functional Signatures in Lymphoid and Mucosal Sites. Cell Rep. 2017;20(12):2921–2934. doi:10.1016/j.celrep.2017.08.078

28. Schapiro D, Jackson HW, Raghuraman S, et al. histoCAT: analysis of cell phenotypes and interactions in multiplex image cytometry data. Nat Methods. 2017;14(9):873–876. doi:10.1038/nmeth.4391

29. Sasson SC, Zaunders JJ, Nahar K, et al. Mucosal-associated invariant T (MAIT) cells are activated in the gastrointestinal tissue of patients with combination ipilimumab and nivolumab therapy-related colitis in a pathology distinct from ulcerative colitis. Clin Exp Immunol. 2020;202:335–352. doi:10.1111/cei.13502

30. van Eijs Mjm, Verheijden RJ, van der Wees SA, et al. Toxicity-specific peripheral blood T and B cell dynamics in anti-PD-1 and combined immune checkpoint inhibition. medRxiv. Published online January 20, 2023. doi:10.1101/2023.01.20.23284818

31. Edelblum KL, Shen L, Weber CR, et al. Dynamic migration of γδ intraepithelial lymphocytes requires occludin. Proc Natl Acad Sci U S A. 2012;109(18):7097–7102. doi:10.1073/pnas.1112519109

32. Corridoni D, Antanaviciute A, Gupta T, et al. Single-cell atlas of colonic CD8+ T cells in ulcerative colitis. Nat Med. 2020;26(9):1480–1490. doi:10.1038/s41591-020-1003-4

33. FitzPatrick MEB, Provine NM, Garner LC, et al. Human intestinal tissue-resident memory T cells comprise transcriptionally and functionally distinct subsets. Cell Rep. 2021;34(3):108661. doi:10.1016/j.celrep.2020.108661

34. Bartolomé-Casado R, Landsverk OJB, Chauhan SK, et al. Resident memory CD8 T cells persist for years in human small intestine. J Exp Med. 2019;216(10):2412–2426. doi:10.1084/jem.20190414

35. Bottois H, Ngollo M, Hammoudi N, et al. KLRG1 and CD103 Expressions Define Distinct Intestinal Tissue-Resident Memory CD8 T Cell Subsets Modulated in Crohn’s Disease. Front Immunol. 2020;11. doi:10.3389/fimmu.2020.00896

36. Dai B, Hackney JA, Ichikawa R, et al. Dual targeting of lymphocyte homing and retention through α4β7 and αEβ7 inhibition in inflammatory bowel disease. Cell Rep Med. 2021;2(8):100381. doi:10.1016/j.xcrm.2021.100381

37. Attauabi M, Madsen GR, Bendtsen F, Seidelin JB, Burisch J. Vedolizumab as the first line of biologic therapy for ulcerative colitis and Crohn’s disease – a systematic review with meta-analysis. Digestive and Liver Disease. 2022;54(9):1168–1178. doi:10.1016/j.dld.2021.11.014

38. Sandborn WJ, Panés J, Danese S, et al. Etrolizumab as induction and maintenance therapy in patients with moderately to severely active Crohn’s disease (BERGAMOT): a randomised, placebo-controlled, double-blind, phase 3 trial. Lancet Gastroenterol Hepatol. 2023;8(1):43–55. doi:10.1016/S2468-1253(22)00303-X

39. Danese S, Colombel JF, Lukas M, et al. Etrolizumab versus infliximab for the treatment of moderately to severely active ulcerative colitis (GARDENIA): a randomised, double-blind, double-dummy, phase 3 study. Lancet Gastroenterol Hepatol. 2022;7(2):118–127. doi:10.1016/S2468-1253(21)00294-6

40. Brummel K, Eerkens AL, de Bruyn M, Nijman HW. Tumour-infiltrating lymphocytes: from prognosis to treatment selection. Br J Cancer. 2023;128(3):451–458. doi:10.1038/s41416-022-02119-4

41. Lucca LE, Axisa PP, Lu B, et al. Circulating clonally expanded T cells reflect functions of tumor-infiltrating T cells. Journal of Experimental Medicine. 2021;218(4). doi:10.1084/jem.20200921

42. Duhen T, Duhen R, Montler R, et al. Co-expression of CD39 and CD103 identifies tumor-reactive CD8 T cells in human solid tumors. Nat Commun. 2018;9(1):2724. doi:10.1038/s41467-018-05072-0

43. Reschke R, Shapiro JW, Yu J, et al. Checkpoint Blockade–Induced Dermatitis and Colitis Are Dominated by Tissue-Resident Memory T Cells and Th1/Tc1 Cytokines. Cancer Immunol Res. 2022;10(10):1167–1174. doi:10.1158/2326-6066.CIR-22-0362

44. Bishu S, Melia J, Sharfman W, Lao CD, Fecher LA, Higgins PDR. Efficacy and Outcome of Tofacitinib in Immune checkpoint Inhibitor Colitis. Gastroenterology. 2021;160(3):932-934.e3. doi:10.1053/j.gastro.2020.10.029

45. Kim ST, Chu Y, Misoi M, et al. Distinct molecular and immune hallmarks of inflammatory arthritis induced by immune checkpoint inhibitors for cancer therapy. Nat Commun. 2022;13(1):1970. doi:10.1038/s41467-022-29539-3

46. Pinal-Fernandez I, Quintana A, Milisenda JC, et al. Transcriptomic profiling reveals distinct subsets of immune checkpoint inhibitor induced myositis. Ann Rheum Dis. Published online February 17, 2023. doi:10.1136/ard-2022-223792

47. Mills KHG. IL-17 and IL-17-producing cells in protection versus pathology. Nat Rev Immunol. 2023;23(1):38–54. doi:10.1038/s41577-022-00746-9

48. Lee JS, Tato CM, Joyce-Shaikh B, et al. Interleukin-23-Independent IL-17 Production Regulates Intestinal Epithelial Permeability. Immunity. 2015;43(4):727–738. doi:10.1016/j.immuni.2015.09.003

49. Schroeder KW, Tremaine WJ, Ilstrup DM. Coated oral 5-aminosalicylic acid therapy for mildly to moderately active ulcerative colitis. A randomized study. N Engl J Med. 1987;317(26):1625–1629. doi:10.1056/NEJM198712243172603

50. NCI (National Cancer Institute (NCI). Common Terminology Criteria for Adverse Events (CTCAE) v5.0. https://ctep.cancer.gov/protocoldevelopment/electronic_applications/ctc.htm#ctc_50.

51. Geboes K, Riddell R, Öst A, Jensfelt B, Persson T, Löfberg R. A reproducible grading scale for histological assessment of inflammation in ulcerative colitis. Gut. 2000;47(3):404–409. http://www.ncbi.nlm.nih.gov/pubmed/10940279 http://www.ncbi.nlm.nih.gov/pubmed/10940279

52. Ma C, MacDonald JK, Nguyen TM, et al. Systematic review: disease activity indices for immune checkpoint inhibitor-associated enterocolitis. Aliment Pharmacol Ther. 2022;55(2):178–190. doi:10.1111/apt.16718

53. Mosli MH, Feagan BG, Zou G, et al. Development and validation of a histological index for UC. Gut. 2017;66(1):50–58. doi:10.1136/gutjnl-2015-310393

54. Forster B, van de Ville D, Berent J, Sage D, Unser M. Complex wavelets for extended depth-of-field: A new method for the fusion of multichannel microscopy images. Microsc Res Tech. 2004;65(1-2):33-42. doi:10.1002/jemt.20092

55. Chalfoun J, Majurski M, Blattner T, et al. MIST: Accurate and Scalable Microscopy Image Stitching Tool with Stage Modeling and Error Minimization. Sci Rep. 2017;7(1):4988. doi:10.1038/s41598-017-04567-y

56. Schindelin J, Arganda-Carreras I, Frise E, et al. Fiji: an open-source platform for biological-image analysis. Nat Methods. 2012;9(7):676–682. doi:10.1038/nmeth.2019

57. Berg S, Kutra D, Kroeger T, et al. ilastik: interactive machine learning for (bio)image analysis. Nat Methods. 2019;16(12):1226–1232. doi:10.1038/s41592-019-0582-9

58. Carpenter AE, Jones TR, Lamprecht MR, et al. CellProfiler: image analysis software for identifying and quantifying cell phenotypes. Genome Biol. 2006;7(10):R100. doi:10.1186/gb-2006-7-10-r100

59. Pebesma E. Simple Features for R: Standardized Support for Spatial Vector Data. R J. 2018;10(1):439. doi:10.32614/RJ-2018-009

60. McInnes L, Healy J, Melville J. UMAP: Uniform Manifold Approximation and Projection for Dimension Reduction. arXiv.

61. Assarsson E, Lundberg M, Holmquist G, et al. Homogenous 96-plex PEA immunoassay exhibiting high sensitivity, specificity, and excellent scalability. PLoS One. 2014;9(4):e95192. doi:10.1371/journal.pone.0095192

62. Hao Y, Hao S, Andersen-Nissen E, et al. Integrated analysis of multimodal single-cell data. Cell. 2021;184(13):3573-3587.e29. doi:10.1016/j.cell.2021.04.048

63. Ahlmann-Eltze C, Huber W. glmGamPoi: fitting Gamma-Poisson generalized linear models on single cell count data. Bioinformatics. 2021;36(24):5701–5702. doi:10.1093/bioinformatics/btaa1009

