## Supplementary material for "Highly multiplexed spatial analysis identifies tissue-resident memory T cells as drivers of ulcerative and immune checkpoint inhibitor induced colitis"

### Supplementary Tables

#### Supplementary Table 1. Patient characteristics

| | IMC cohort (colitis) | | | | Serum cohort (irAEs including colitis) | | | |
|---|---|---|---|---|---|---|---|---|
| | | | | | αPD-1 | | cICI | |
| | **αCTLA-4** (n=4) | **αPD-1** (n=5) | **cICI** (n=9) | **UC** (n=5) | **irAE+** (n=15) | **irAE-** (n=25) | **irAE+** (n=24) | **irAE-** (n=16) |
| **Age**, median (p25–p75), *yr* | 75 (67-77) | 71 (67-72) | 67 (51-72) | 51 (36-52) | 73 (65-75) | 63 (58-73) | 55 (48-67) | 71 (53-77) |
| **Sex**, *N* female (%) | 1 (25) | 3 (60) | 4 (44) | 1 (20) | 5 (33) | 5 (20) | 8 (33) | 7 (44) |
| **Prednisone duration (pre-endoscopy)**, median (p25–p75), *days* | 1 (0-3) | 0 (0-0) | 1 (0-5) | 0 (0-0) | NA | NA | NA | NA |
| **Symptom duration (pre-endoscopy)**, median (p25–p75), *days* | 8 (4-12) | 14 (12-38) | 7 (6-11) | 60 (40-90) | NA | NA | NA | NA |
| **Mayo endoscopic score**, *N* (%) | | | | | NA | NA | NA | NA |
| 0 | 0 (0) | 0 (0) | 1 (11) | 0 (0) | | | | |
| 1 | 1 (25) | 4 (80) | 4 (44) | 1 (20) | | | | |
| 2 | 2 (50) | 1 (20) | 2 (22) | 3 (60) | | | | |
| 3 | 1 (25) | 0 (0) | 0 (0) | 1 (20) | | | | |
| **New-onset ulcerative colitis**, *N* (%) | NA | NA | NA | 3 (60) | NA | NA | NA | NA |
| **Concomitant irAEs besides main irAE***, *N* (%) | | | | NA | | NA | | NA |
| 0 | 4 (100) | 2 (40) | 4 (44) | | 13 (87) | | 19 (79) | |
| 1 | 0 (0) | 3 (60) | 2 (22) | | 1 (7) | | 2 (8) | |
| 2 | 0 (0) | 0 (0) | 2 (22) | | 1 (7) | | 1 (4) | |
| >2 | 0 (0) | 0 (0) | 1 (11) | | 0 (0) | | 2 (8) | |
| **Colitis among irAEs**, *N* (%) | 4 (100) | 5 (100) | 9 (100) | NA | 3 (20) | NA | 11 (46) | NA |
| **Systemic immuno-suppression required post-endoscopy**, *N* (%) | | | | | NA | NA | NA | NA |
| None (only mesalazine +/- topical IS) | 0 (0) | 0 (0) | 0 (0) | 3 (60) | | | | |
| Steroids only | 0 (0) | 2 (40) | 3 (33) | 1 (20) | | | | |
| Steroids + anti-TNF | 4 (100) | 3 (60) | 6 (67) | 0 (0) | | | | |
| Steroids + anti-TNF + azathioprine | 0 (0) | 0 (0) | 0 (0) | 1 (20) | | | | |

* 'main' irAE indicates ICI-colitis in IMC cohort, or clinically most relevant/most severe irAE in Serum cohort. αCTLA-4 denotes 'anti-CTLA-4 monotherapy', αPD-1 'anti-PD-1 monotherapy', cICI 'combined anti-CTLA-4 and anti-PD-1', irAE 'immune-related adverse event', IS 'immunosuppression', NA 'not applicable', UC 'ulcerative colitis', yr 'years'

**Supplementary Table 2. Antibodies and channels used for imaging mass cytometry**

| Isotope | Target | Clone | Dilution | Company | Catalogue no. |
|---|---|---|---|---|---|
| Nd-142 | E-cadherin | 24E10 | 100 | Cell Signaling Technology | CST3195BF |
| Nd-143 | CD68 | KP1 | 600 | Thermo Fisher Scientific | 14-0688-82 |
| Nd-144 | CD14 | EPR3653 | 400 | Abcam | ab214438 |
| Nd-145 | pS6 | D57.2.2E | 100 | Cell Signaling Technology | CST4858BF |
| Nd-146 | CD20 | H1 | 150 | BD Biosciences | 555677 |
| Sm-147 | CD45RA | HI100 | 100 | Biolegend | 304102 |
| Nd-148 | HLA-DR+DP+DQ | CR3/43 | 200 | Abcam | ab7856 |
| Sm-149 | CTLA-4 | UMAB249 | 100 | OriGene Technologies | UM800141CF |
| Nd-150 | RORγt | RORC/2941 | 50 | Abcam | ab268233 |
| Eu-151 | IFNγ | D3H2 | 100 | Cell Signaling Technology | CST8455BF |
| Sm-152 | CD45RO | UCHL1 | 100 | Cell Signaling Technology | CST55618BF |
| Eu-153 | CD103 | EPR4166(2) | 200 | Abcam | ab221210 |
| Sm-154 | TNFα | 7B8A11 | 50 | Proteintech | 60291-1-1g |
| Gd-155 | FoxP3 | 236A/E7 | 50 | Abcam | ab96048 |
| Gd-156 | CD4 | EPR6855 | 50 | Abcam | ab181724 |
| Gd-158 | T-bet | D6N8B | 50 | Cell Signalling Technology | CST13232BF |
| Tb-159 | CD45 | D9M8I | 100 | Cell Signaling Technology | CST13917BF |
| Gd-160 | IL-10 | polyclonal | 150 | R&D Systems | AF-217-NA |
| Dy-161 | TIGIT | polyclonal | 50 | Abcam | ab233404 |
| Dy-162 | CD8α | C8/144B | 200 | Thermo Fisher Scientific | 14-0085-82 |
| Dy-163 | ICOS | D1K2T | 100 | Cell Signaling Technology | CST89601BF |
| Ho-165 | PD-1 | EPR4877(2) | 50 | Abcam | ab186928 |
| Er-166 | CD56 | NCAM1/784 | 200 | Abcam | ab216010 |
| Er-167 | IL-17A | polyclonal | 100 | R&D Systems | AF-317-NA |
| Er-168 | Ki67 | B56 | 200 | BD Biosciences | 556003 |
| Tm-169 | Granzyme B | D6E9W | 100 | Cell Signaling Technology | CST46890BF |
| Er-170 | CD3 | polyclonal | 100 | Dako | A045229-2 |
| Yb-171 | CD11c | EP1347Y | 100 | Abcam | ab216655 |
| Yb-172 | IL12Rβ2 | polyclonal | 50 | Thermo Fisher Scientific | PA5-24181 |
| Yb-173 | CD69 | polyclonal | 150 | Proteintech | 10803-1-AP |
| Yb-174 | TCR-δ | H-41 | 100 | Santa Cruz Biotechnology | sc-100289X |
| Yb-176 | Histone H3 | D1H2 | 600 | Cell Signaling Technology | CST4499BF |

### Supplementary Figures

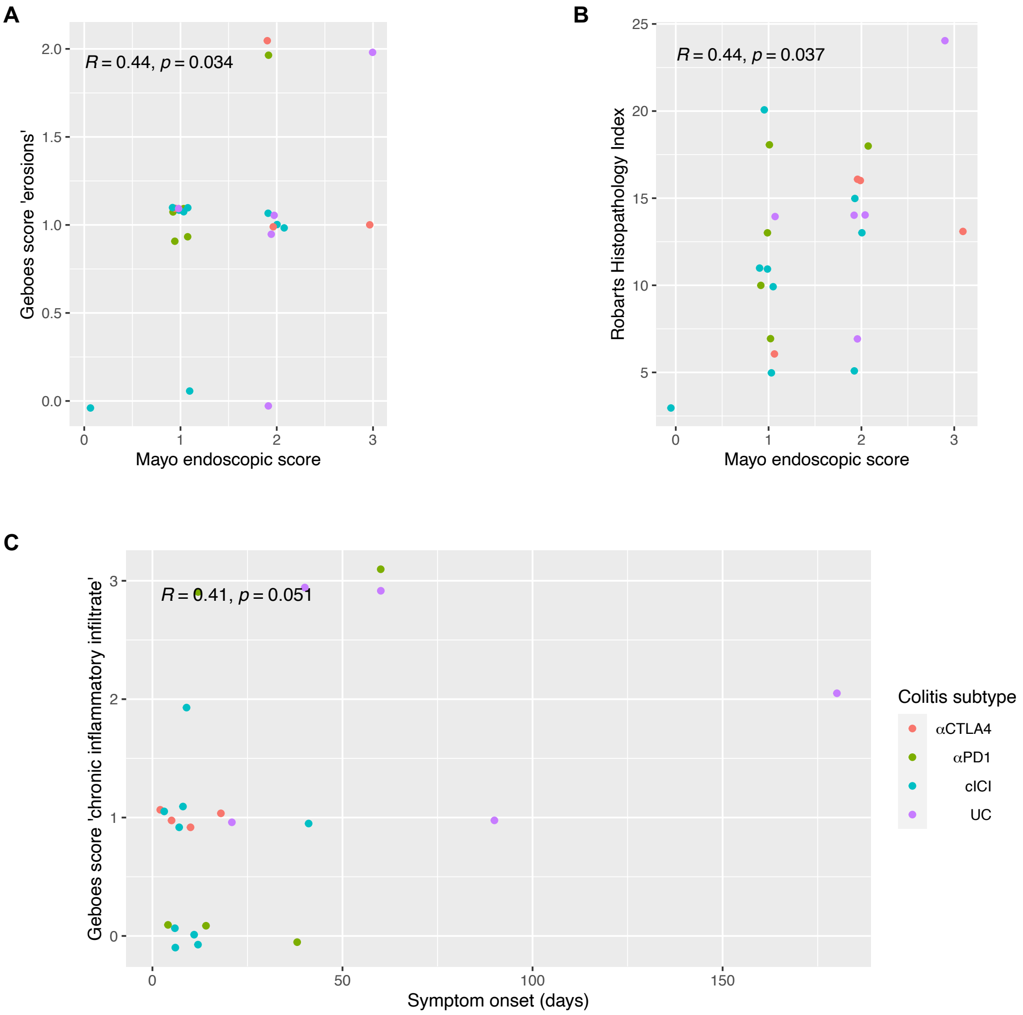


**Supplementary Figure 1. Correlation of clinical colitis severity indices within the IMC cohort.** (**A**-**C**) Associations between (**A**) Geboes score for erosions and Mayo endoscopic score, (**B**) Robarts Histopathology Index and Mayo endoscopic score and (**C**) Geboes score for chronic inflammatory infiltrate and duration of symptoms, with maximum 0.1 horizontal and vertical jitter to prevent overplotting. *R* denotes Spearman's rank correlation coefficient (n=23 patients).

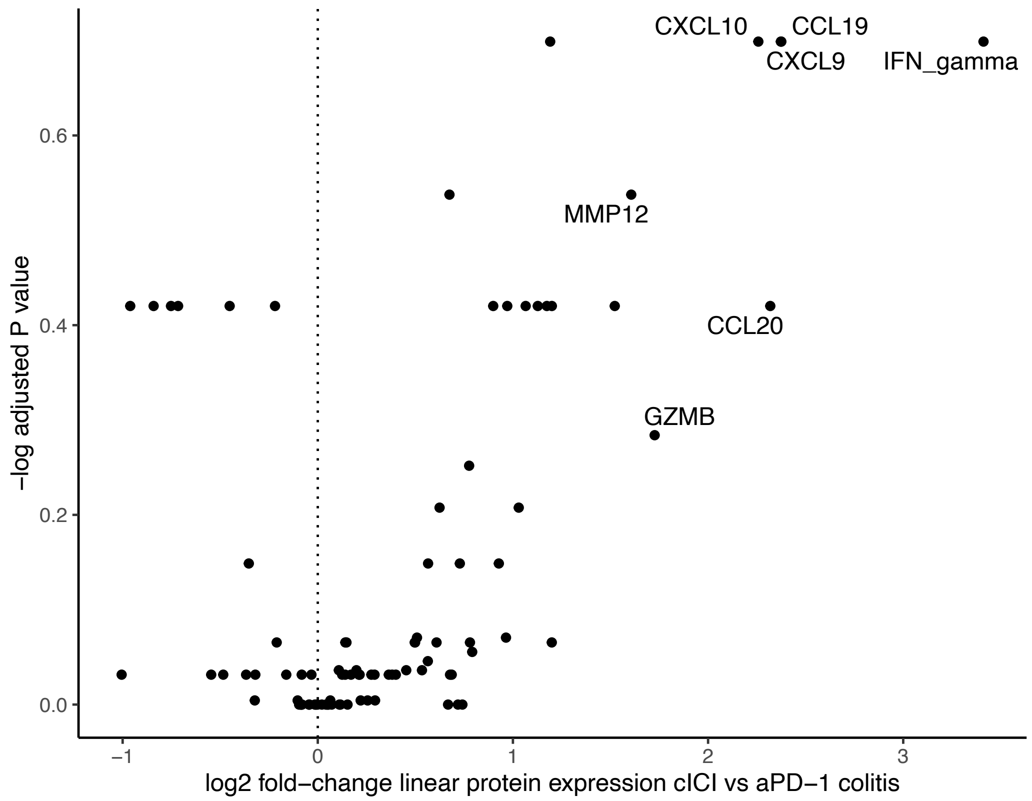


**Supplementary Figure 2. Differential serum protein expression for cICI versus αPD-1 colitis.**

Volcano plot showing differential serum protein expression of 92 analytes in colitis (with or without concomitant other irAEs) after combined αCTLA-4 and αPD-1 relative to αPD-1 monotherapy (only samples obtained upon colitis onset, n=14). Differential expression analyzed by Wilcoxon test with Benjamini-Hochberg false-discovery rate correction.

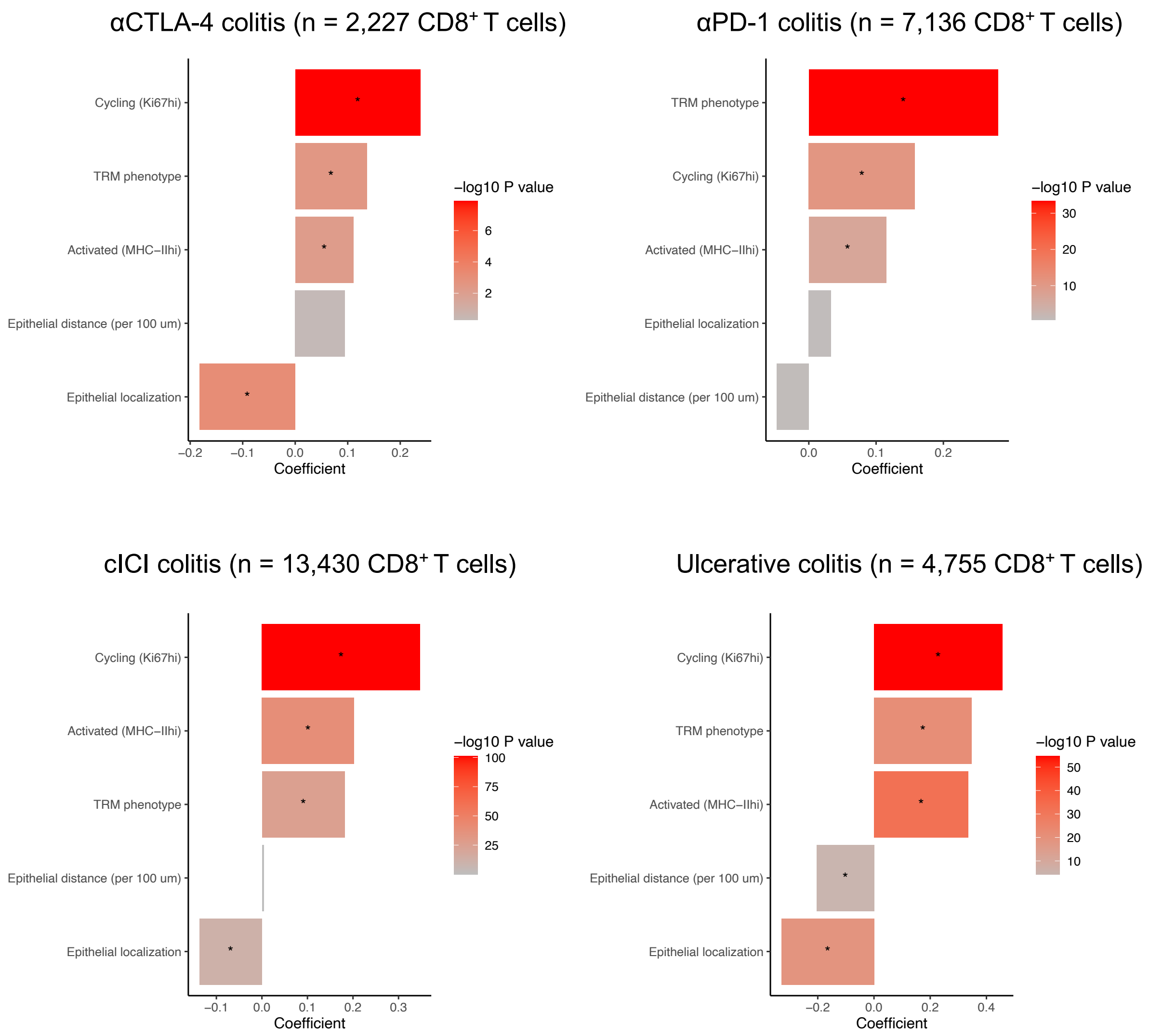


**Supplementary Figure 3. Analysis of factors contributing to CD8$^+$ granzyme B production stratified by colitis subtype**

Visualization of coefficients and -log$_{10}$(*P* values) of fixed effects. Mixed-effects models with fixed effects for all listed covariates and random intercepts for individual patients were fit to explain CD8$^+$ T cell granzyme B production, stratified by colitis subtype. Coefficients >0.0 indicate that CD8$^+$ granzyme B levels are higher in presence of (or for higher values of) the variable. *$P$<0.05.

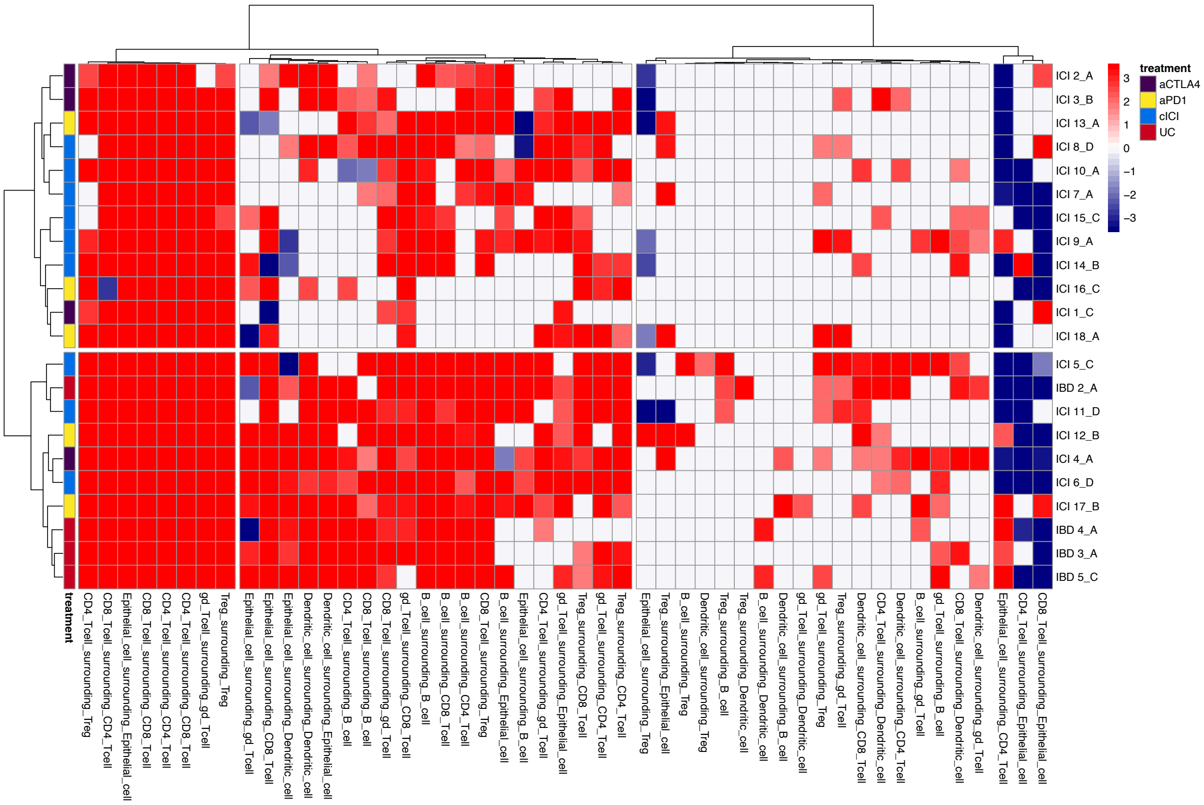


**Supplementary Figure 4. Agglomerative clustering of cell-cell interactions by neighborhood analysis.**

Heatmap showing Benjamini-Hochberg false-discovery rate corrected “directional” $-\log_{10} P$ values of interactions among epithelial cells, CD4$^+$ and CD8$^+$ T cells, CD4$^+$ Tregs, dendritic cells, B cells and $\gamma\delta$ T cells. Significant interactions are annotated in red, meaning that two cell types are close to each other more frequently than expected by chance. Significant avoidance is annotated in blue.

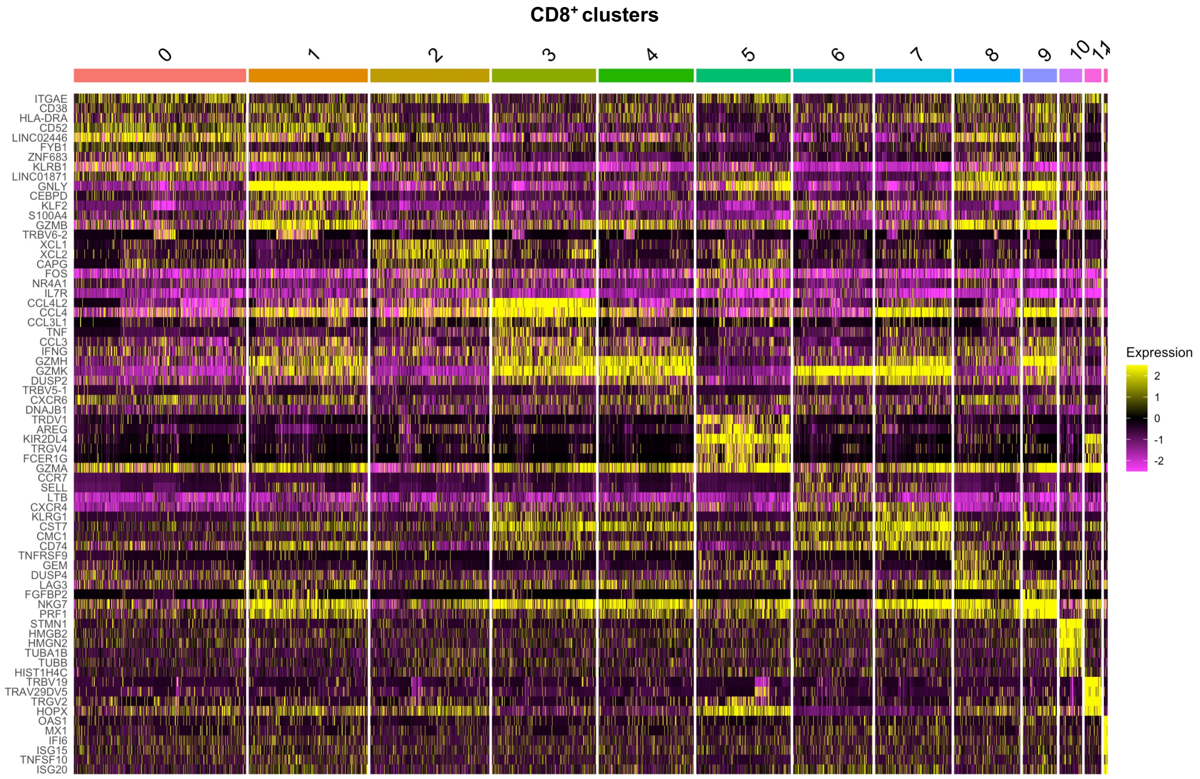


**Supplementary Figure 5. CD8$^+$ cluster-defining feature heatmap.**

Expression heatmap showing top 6 differentially expressed genes per CD8$^+$ cell cluster along with ITGAE, CD38 and HLA-DRA (top 3 rows).

A

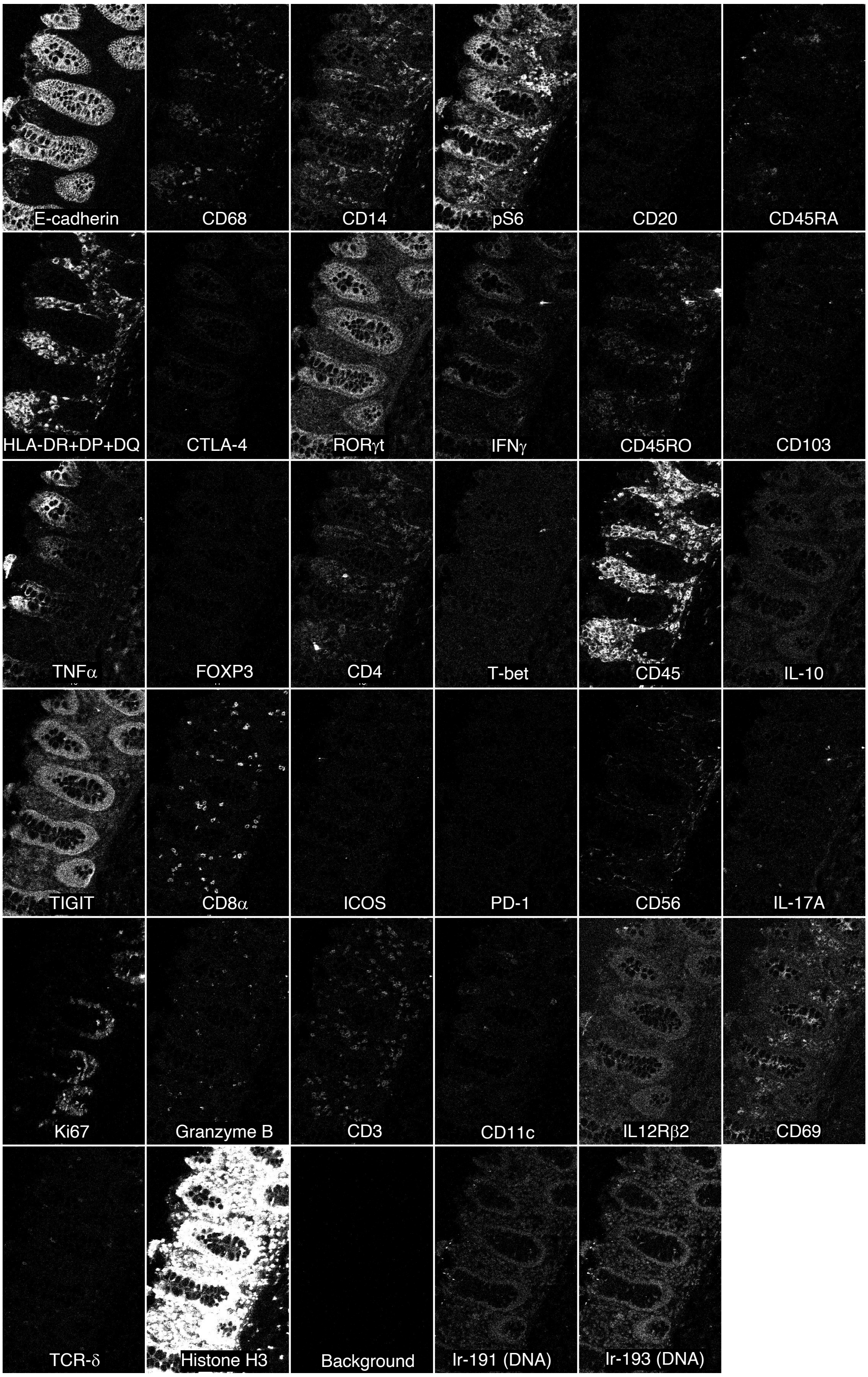
E-cadherin
CD68
CD14
pS6
CD20
CD45RA
HLA-DR+DP+DQ
CTLA-4
RORγt
IFNγ
CD45RO
CD103
TNFα
FOXP3
CD4
T-bet
CD45
IL-10
TIGIT
CD8α
ICOS
PD-1
CD56
IL-17A
Ki67
Granzyme B
CD3
CD11c
IL12Rβ2
CD69
TCR-δ
Histone H3
Background
Ir-191 (DNA)
Ir-193 (DNA)

B

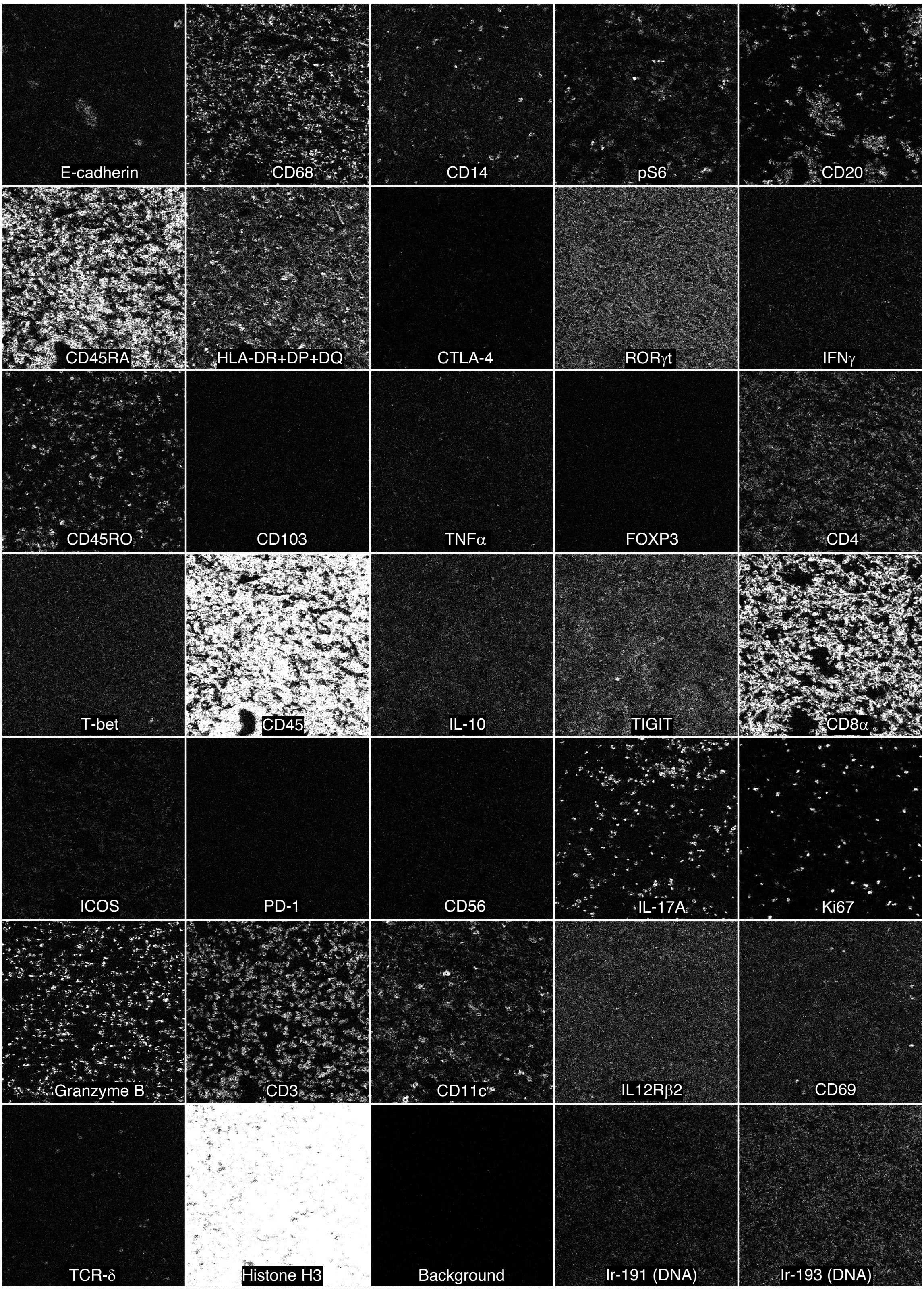


**Supplementary Figure 6. IMC single channel expression data in tissue microarray controls**

(**A**,**B**) Antigen expression in all channels is displayed for (**A**) uninflamed colon and (**B**) ovarian adenocarcinoma tissue, included as tissue microarray on all slides used for IMC.

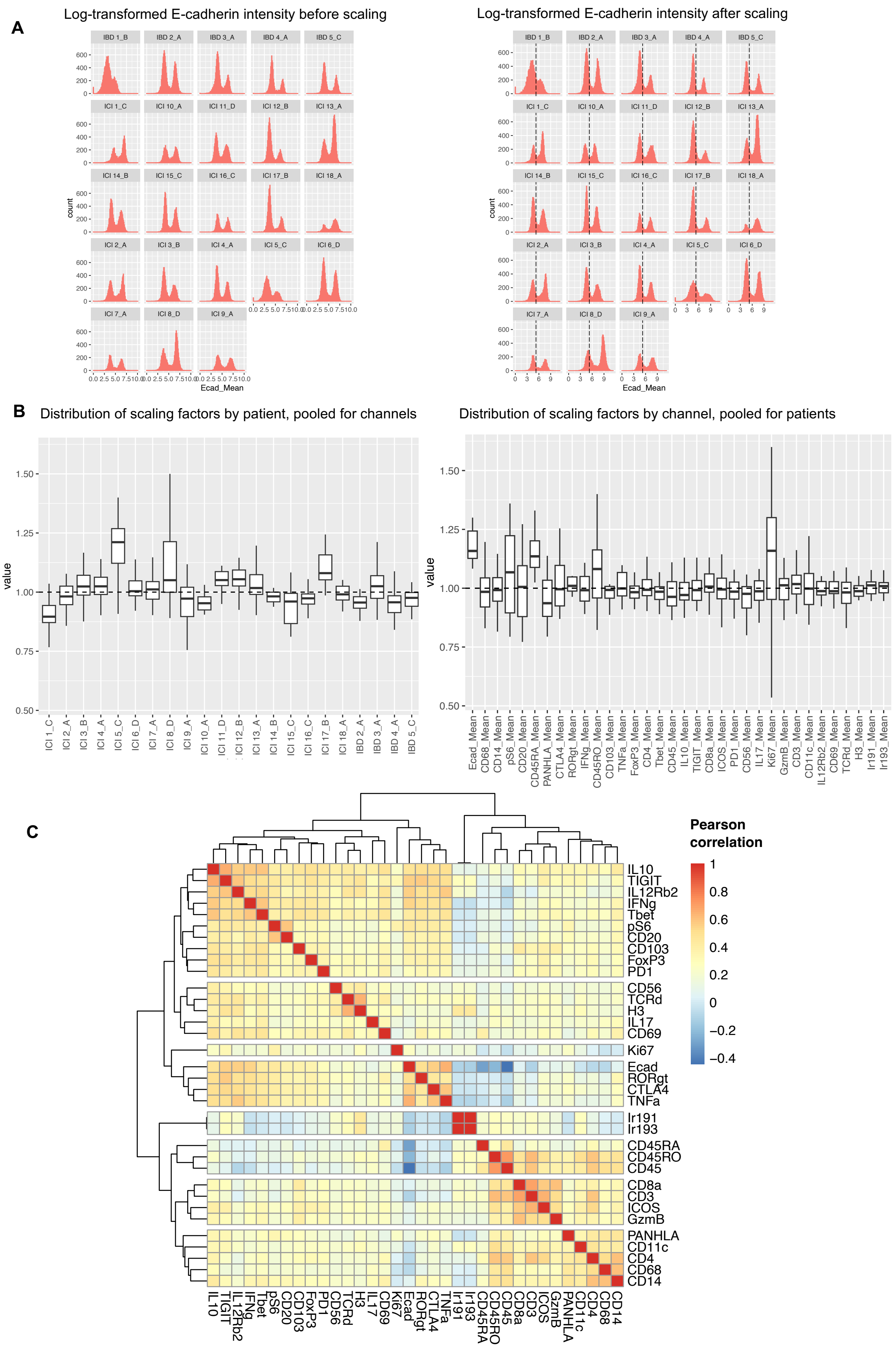


**Supplementary Figure 7. IMC data scaling and marker co-expression.**

(**A**) Patient-specific histograms of natural-log transformed E-cadherin signal before (left) and after (right) scaling. The goal of scaling was to align channel expression data in such way that (1) a single threshold for marker positivity could be used across patients and (2) marker expression levels could be directly compared between patients. Note that the dashed line, indicating marker positivity threshold for E-cadherin, is intentionally set at a too low level. In this way, the number of false negative cells (expressing E-cadherin but falsely labeled as E-cadherin negative) that would not proceed to rank-based lineage determination is minimized.

(**B**) Boxplots of linear scaling factors used. Values >1 indicate that the original intensities were numerically increased (visually histograms as in (**A**) would shift to the right).

(C) Heatmap showing correlation (by Pearson's correlation coefficient) of cellular expression among all markers. This confirms that markers showing highly correlated expression are also biologically associated, e.g., CD3, CD8α, granzyme B (GzmB) and ICOS. Considerable non-specific binding of CTLA-4, RORγt and potentially TNF-α antibodies to epithelium was suspected, as supported by this figure, and therefore these markers were not used in further analysis.

**Annotated pseudocolor plot IBD 1_B**

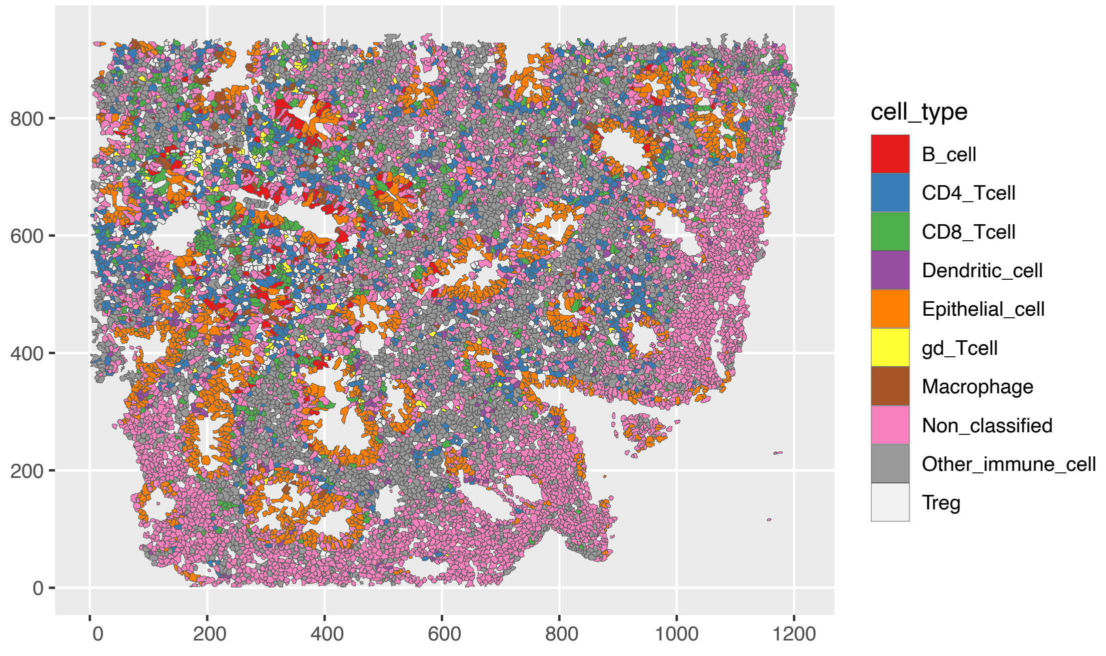


**Raw IMC image IBD 1_B**

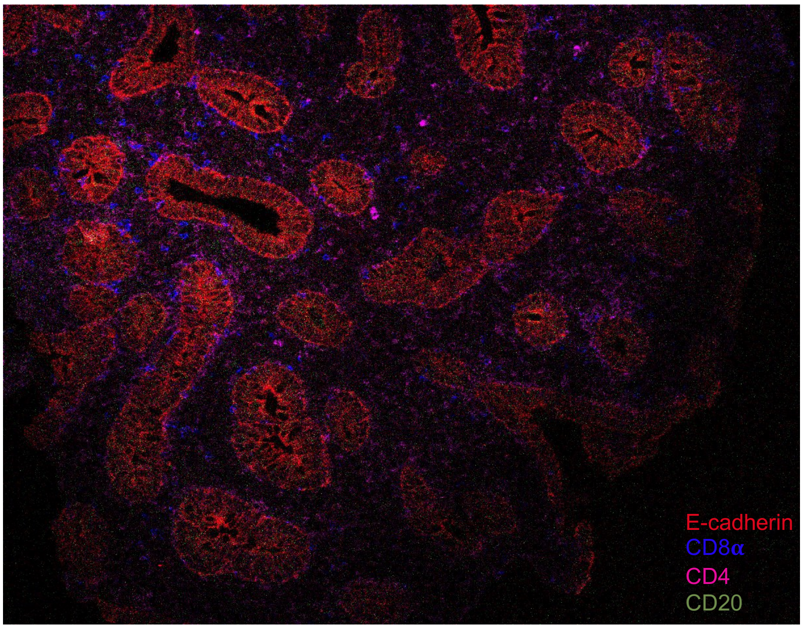


**Supplementary Figure 8. Low quality of IMC data in IBD 1_B.**

Pseudo-color plot (top) showing that the majority of cells was either non-classified (37%), or classified as other immune cells (27%). This was the result of large within-image variation in signal intensity (bottom) that could not properly be improved, which led to the decision to exclude IBD 1_B from further analysis.
